# Prevalence of Mental Disorders in Parents with Intellectual Disabilities: Systematic Review and Meta-Analysis

**DOI:** 10.1101/2025.10.09.25337705

**Authors:** Sonya Rudra, Louise Marston, Angela Hassiotis, Giovanni Boido, Mary Baginsky, Claire A Wilson

**Affiliations:** Division of Psychiatry, University College London, London, UK; Department of Primary Care and Population Health, University College London, London, UK; Department of Health Studies, University of Milan, Milan, Italy; Department of Mental Health and Addiction - ASST Santi Paolo e Carlo, Milan, Italy; NIHR Health and Social Care Workforce Research Unit, Policy Institute, King’s College London, UK; South London and Maudsley NHS Foundation Trust, London, UK; Institute of Psychiatry, Psychology and Neuroscience, Kings College London, UK

## Abstract

**Background:** Research indicates that increasing numbers of people with intellectual disabilities (ID) are becoming parents. Parents with ID face multiple contextual factors that elevate mental health risks during the perinatal period. Mental disorders in this period can significantly impact parents, their children, and the wider support network.

**Aim:** This systematic review summarises the prevalence of mental disorders in parents with ID during pregnancy, post-partum, and parenthood, comparing rates with the general population and non-parents with ID.

**Methods:** Studies included individuals with clinically diagnosed ID who had experienced pregnancy or parenthood and reported prevalence of specific mental disorders. Searches of MEDLINE, APA PsycINFO, EMBASE, and grey literature were conducted to May 2025. Risk of bias was assessed using the Mixed Methods Appraisal Tool. Findings were synthesised narratively, with meta-analysis where possible.

**Results:** Ten studies involving 42,207 individuals aged 18–58 years were included. Two studies focused on the perinatal period, the rest on parents with children aged 0–21 years. Across studies, approximately half of parents with ID experienced a psychiatric disorder. Depression was most common, affecting 38% (95% CI: 26– 50%) mothers, anxiety 26% (95% CI: 26-26%), and psychosis 9% (95% CI: 3–14%). Rates are higher than in the general population and non-parent individuals with ID.

**Conclusion:** This review is the first to document elevated mental disorders among parents with ID. Findings highlight the need for tailored screening and support in reproductive healthcare, addressing stigma, and guiding further research to inform targeted interventions.

PROSPERO registration number 1020203

**What this paper adds?:** This review is the first to systematically synthesise evidence demonstrating elevated risks of mental disorders among parents with intellectual disabilities (ID), showing that around half of parents experience a psychiatric disorder, with depression affecting 38% of mothers, anxiety 26%, and psychosis 9% — rates higher than in the general population and non-parent individuals with ID. It highlights the substantial mental health burden faced by this group and identifies major gaps in the literature, including limited research on fathers, postpartum mental health, studies based on larger samples and studies from low- and middle-income countries. In doing so, this paper provides a foundation for future research and highlights the need for targeted, evidence-based interventions and tailored mental health screening and support for parents with ID, particularly during the perinatal period.

**Highlights:**

- There are high rates of mental disorders in parents with intellectual disabilities
- Half of parents with intellectual disabilities have psychiatric disorders
- Research gaps exist on postpartum health and fathers with intellectual disabilities
- There is urgent need for tailored mental healthcare in reproductive health services

## 1. INTRODUCTION

### 1.1. Defining Intellectual Disability

Intellectual Disability (ID) is a lifelong disorder that affects around 1% of the worldwide population (Maulik et al., 2011). In both the Diagnostic and Statistical Manual of Mental Disorders, Fifth Edition (DSM-5) and International Classification of Diseases, 11th Revision (ICD-11) the core diagnostic criteria for ID (also referred to as “intellectual developmental disorder” in the DSM-5 and “disorders of intellectual development” in the ICD-11 are: deficits in intellectual functioning; deficits in adaptive functioning and; onset during the developmental period (American Psychiatric Association, 2013; World Health Organization, 2022). These criteria place less of an emphasis on IQ scores compared to earlier diagnostic manuals (American Psychiatric Association, 1994; World Health Organization, 2004).

### 1.2. Parenthood Trends in Intellectual Disability

Research suggests that it is likely more people with ID are becoming parents (Turney et al., 2018). This rise in parents can be attributed to a move to more community-based care providing opportunities to develop relationships, coupled with changing attitudes, a reduction in forced sterilization, as well as policy and legislative changes recognising the rights of people with disabilities to parent and to have a family life (Human Rights Act 1998; Equality Act, 2010).

### 1.3. Contextual Risks for Perinatal Mental Disorders in Intellectual Disability

Contextual factors are important in understanding perinatal mental disorders. Prevalence estimates suggest that depression and anxiety are at least as common, if not more so, in the perinatal period compared to other life stages, affecting approximately one in five women globally (Stevenson et al., 2023). In general, people with ID face significant health and social disparities including higher rates of poverty, poor physical and mental health, and exposure to violence compared to those without ID (Bowen & Swift, 2019; Emerson, 2007; Krahn et al., 2006; Roebuck, 2021). These factors culminate in higher rates of avoidable mortality in those with ID compared to the general population (White et al., 2023).

Mothers with ID therefore face a double burden increasing their mental health risk during the perinatal period. Research has suggested that they are more likely to be unemployed, struggle financially, and reside in deprived neighbourhoods compared to mothers without ID (Brown et al., 2022; Fairthorne et al., 2020; Tarasoff et al., 2020). They also tend to be younger (Parish et al., 2015) and have a higher likelihood of being single parents (Mitra et al., 2015), which are both risk factors for isolated parenting experiences and victimization. Women with ID may also suffer high rates of trauma, such as child welfare involvement, intimate partner violence, sexual assault, and caregiver abuse (Bowen & Swift, 2019; Powell et al., 2024), often experiencing these life events with smaller support networks and without appropriately tailored support (Mueller et al., 2019)

### 1.4. Pregnancy and Postnatal Outcomes in Women with Intellectual Disability

A recent systematic review examining pregnancy and postnatal outcomes of women with ID found that women with ID were at an overall higher risk of adverse obstetric and pregnancy outcomes, such as gestational hypertension, and postpartum haemorrhage compared to women without ID (Lo et al., 2025). Similarly, infants of women with ID had higher rates of premature birth, perinatal mortality, and experienced longer hospital stays when compared to their counterparts born to women without ID (Lo et al., 2025).

Pregnancy outcomes are worse still for mothers with comorbid ID and mental illness (Brown et al., 2016). A complex cycle exists in which adverse experiences and poor physical outcomes increase the risk of maternal mental health issues (Anderson & Cacola, 2017).

### 1.5. Impact of Maternal Mental Disorders in Intellectual Disability

Mental disorders in parents with ID can have a significant impact on the parent, their children, and the wider network. For women, it can impact their ability to effectively parent, interact with their children and enjoy motherhood (Ehlers-Flint, 2002). Mental disorders in this group are frequently cited as a contributory factor in the permanent removal of children from their families (Booth et al., 2005). For children, they can become more susceptible to adverse neuro-behavioural outcomes (such as attention deficits, anxiety disorders and autism) than those children whose parents have ID without mental illness (McGaw et al., 2007). For services, during pregnancy and after childbirth, women with ID have higher rates of emergency department visits and hospital admissions for psychiatric reasons including schizophrenia/psychosis, postpartum depression, and bipolar affective disorder (Brown et al., 2017).

### 1.6. Limitations in Research on Mental Disorders in Parents with Intellectual Disability

A number of studies have used administrative health data to show that parents with “intellectual and developmental disabilities” (IDD) have higher rates of some mental disorders (Brown et al., 2016). However, there is often no separate analysis for parents with ID only. Parents with ID may have different health needs than individuals with neurodevelopmental disorders such as autism without ID. In many countries, as in England, these groups are not eligible for ID services if they do not have comorbid ID. It is therefore imperative for clinicians and professionals working with people with ID to have an awareness of the needs of the population with ID specifically to provide personalised and tailored care.

In some studies which focus on ID, samples are identified by clinical suspicion or self-report rather than formal diagnosis (Munshi et al., 2025). This can introduce bias into the sample, including the inclusion of participants who have “learning difficulties” referring to limitations in scholastic skills rather than a clinical ID diagnosis.

Research focusing on fathers, particularly those with ID, is scarce. Nonetheless, evidence indicates that fathers can experience significant mental health difficulties linked to service exclusion and the compounded stresses of parenthood (Symonds et al., 2021). In the general population, perinatal depression affects 8–26% of fathers, markedly higher than the approximate 5% annual prevalence among men overall (Bruno et al., 2020). This highlights the need to examine whether paternal depression rates in the male ID population follow a similar trend.

### 1.7. Study Objectives

This research will synthesize the existing evidence on mental disorders in parents with ID. The primary objective is to systematically review the literature for evidence of the prevalence of any mental disorders in parents with ID during pregnancy, post-partum, and parenthood. We also aim to comment on whether parents with ID have a higher prevalence of mental disorders compared to the general population and non-parents with ID.

## 2. METHODOLOGY

### 2.1. Eligibility and Outcomes

We included quantitative studies of parents with clinically defined ID (DSM/ICD or equivalent), and partners of pregnant people, without restrictions on country or setting. We excluded selfl7lreport–only ID, algorithmic/composite ID definitions, borderline intellectual functioning, and Intimate Partner Violence/childl7lprotection cohorts. Primary outcomes were prevalence of clinically diagnosed or screened mental disorders during pregnancy, postpartum, or parenthood. Reviews, case studies and opinion pieces were excluded.

### 2.2. Search Strategy

We searched MEDLINE, APA PsycINFO, and EMBASE via OVID from inception to May 2025, plus grey literature (PsycEXTRA, PsyArXiv, ProQuest Dissertations and Theses). The search strategy was developed with a specialist librarian, peerl7lreviewed (PRESS) (McGowan et al., 2016) and validated against known studies. Searches were also done on reference lists from key reviews and to identify corresponding papers from relevant conference abstracts. Duplicates were removed using the Falconer (2018) method, imported into Rayyan for screening and citation tracking completed in Scopus and Web of Science. Database search alerts were set in order to identify any new publications prior to starting the data extraction in July 2025. The search strategy is available in Supplementary material (S1).

### 2.3. Selection Process

Two reviewers independently screened titles/abstracts (initial 20% doublel7lscreened with full agreement) and then full texts against eligibility criteria; nonl7lEnglish abstracts were translated; disagreements were resolved by discussion or a third reviewer.

### 2.4. Data Collection

Two reviewers independently extracted study characteristics, ID definitions, outcomes, and comparator details using a piloted form; authors were contacted for missing data.

### 2.5. Risk of Bias Assessment

The Mixed Methods Appraisal Tool (MMAT; Hong et al. (2018)), was used to assess the risk of bias within studies. This tool evaluates qualitative, quantitative, and mixed methods studies using two universal screening questions and five design-specific criteria, rated as “Yes,” “No,” or “Can’t tell,” to ensure transparent appraisal across study types. The studies were rated by two researchers independently. In order to aid comparisons across studies, a final judgement was given by each assessor depending on whether the study met the criteria on all 5 questions (low risk of bias), 4 questions (moderate risk of bias), or ≤3 questions (high risk of bias). Any missing or unclear data was judged to reduce the overall risk of bias of the paper. Ratings were combined and any disagreements resolved through discussion between the two researchers, involving a third researcher if required.

### 2.6. Data Synthesis

Results are presented in an initial narrative analysis, based on population characteristics, study design and outcomes. Where three or more studies present prevalence data for a specific mental disorder, StataNow MP 19.5 was used to pool studies using random-effects meta-analyses. Statistical heterogeneity was assessed using the I^2^ statistic, representing percentage variation between sample estimates due to heterogeneity.

In a further narrative analysis, the prevalence rates (N, %) of mental disorders were compared to prevalence rates of the same conditions reported in non-parents with ID or in the general population, derived from the same papers or from the wider literature.

Subgroup analyses were planned to compare studies using clinical diagnoses versus screening tools, and studies focused on pregnancy versus postpartum. Publication bias was to be assessed using funnel plots and Egger’s regression tests if ≥10 studies were available. Sensitivity analyses excluding high-risk studies were also planned. These analyses were not conducted due to insufficient data.

## 3. RESULTS

### 3.1. Study Selection

The search yielded 12,957 records; after deduplication, 8,322 were screened, 48 full texts assessed, and ten studies met inclusion criteria. See Supplementary material (S2) for articles excluded. The study selection process is illustrated in **Figure 1: PRISMA Diagram.**

**Figure 1:**
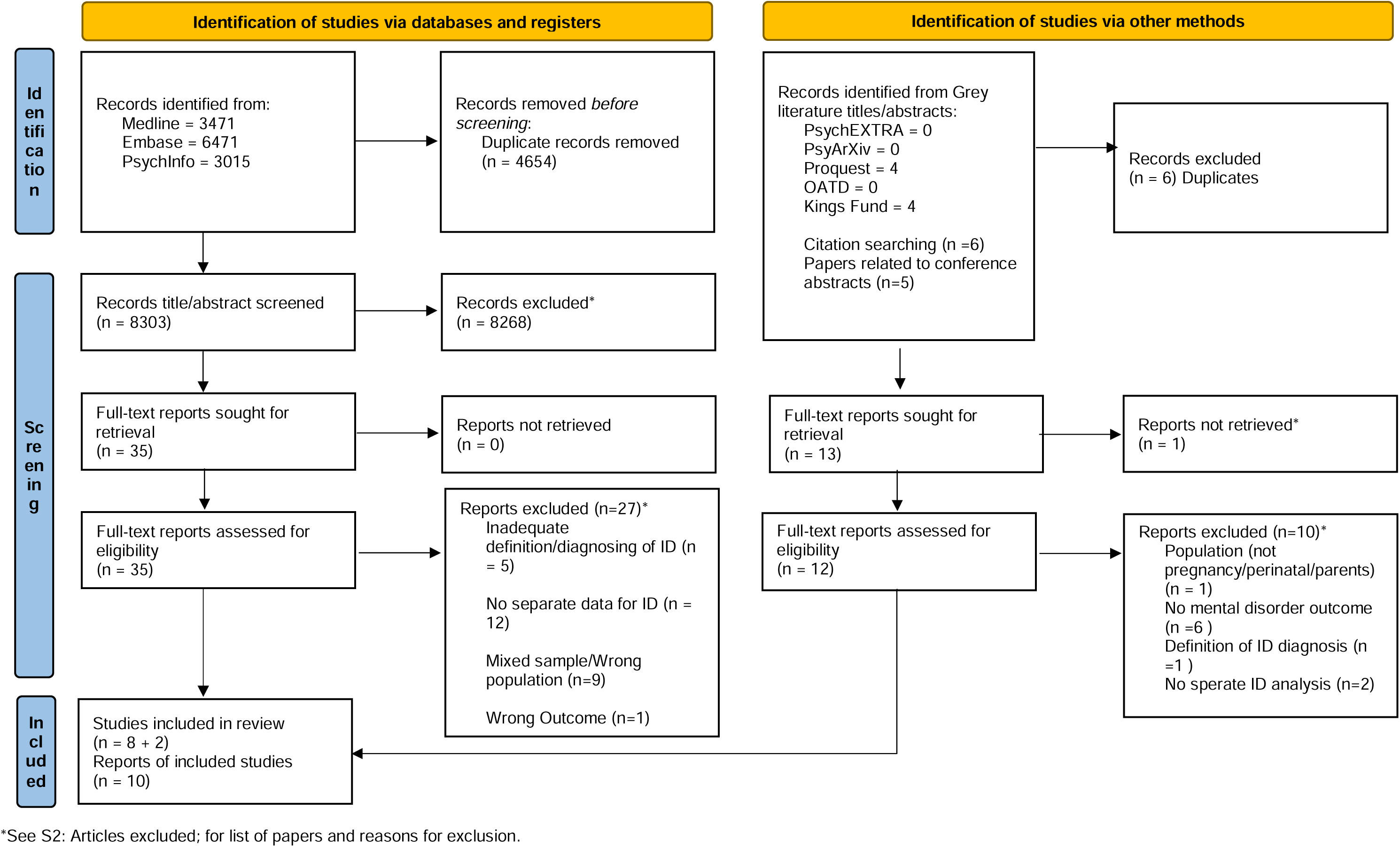
PRISMA flow diagram for systematic reviews which included searches of databases and other sources. The study selection process is illustrated: Initial database search yielded 12,957 records. After duplicates were removed, 8322 records remained for title and abstract review, including 19 papers identified via other methods (grey literature, citation searching and papers related to conference abstracts). Following a review of title/abstracts, 48 articles were found eligible for full-text screening. 37 articles were excluded during full-text screening and one potential full-text based on conference abstract was unable to be retrieved. Overall, 10 studies were eligible and included in this review

### 3.2. Study Characteristics

The search identified a combination of cross-sectional studies (Gaskin & James, 2006; Sterling, 1998; Tymchuk, 1993; Tymchuk, 1994; Walton-Allen, 1993), cohort studies (Lindblad et al., 2024; Shea et al., 2024), retrospective observational studies (McGaw et al., 2010; McGaw et al., 2007) and a qualitative study with quantitative component (Heifetz et al., 2019). Two thesis papers were identified and included in the analysis. All studies identified were from high-income countries: USA (Shea et al., 2024; Sterling, 1998; Tymchuk, 1993; Tymchuk, 1994), UK (Gaskin & James, 2006; McGaw et al., 2010; McGaw et al., 2007), Sweden (Lindblad et al., 2024) and Canada (Heifetz et al., 2019; Walton-Allen, 1993). **Table 1** shows the characteristics of the included studies.

**Table 1:**
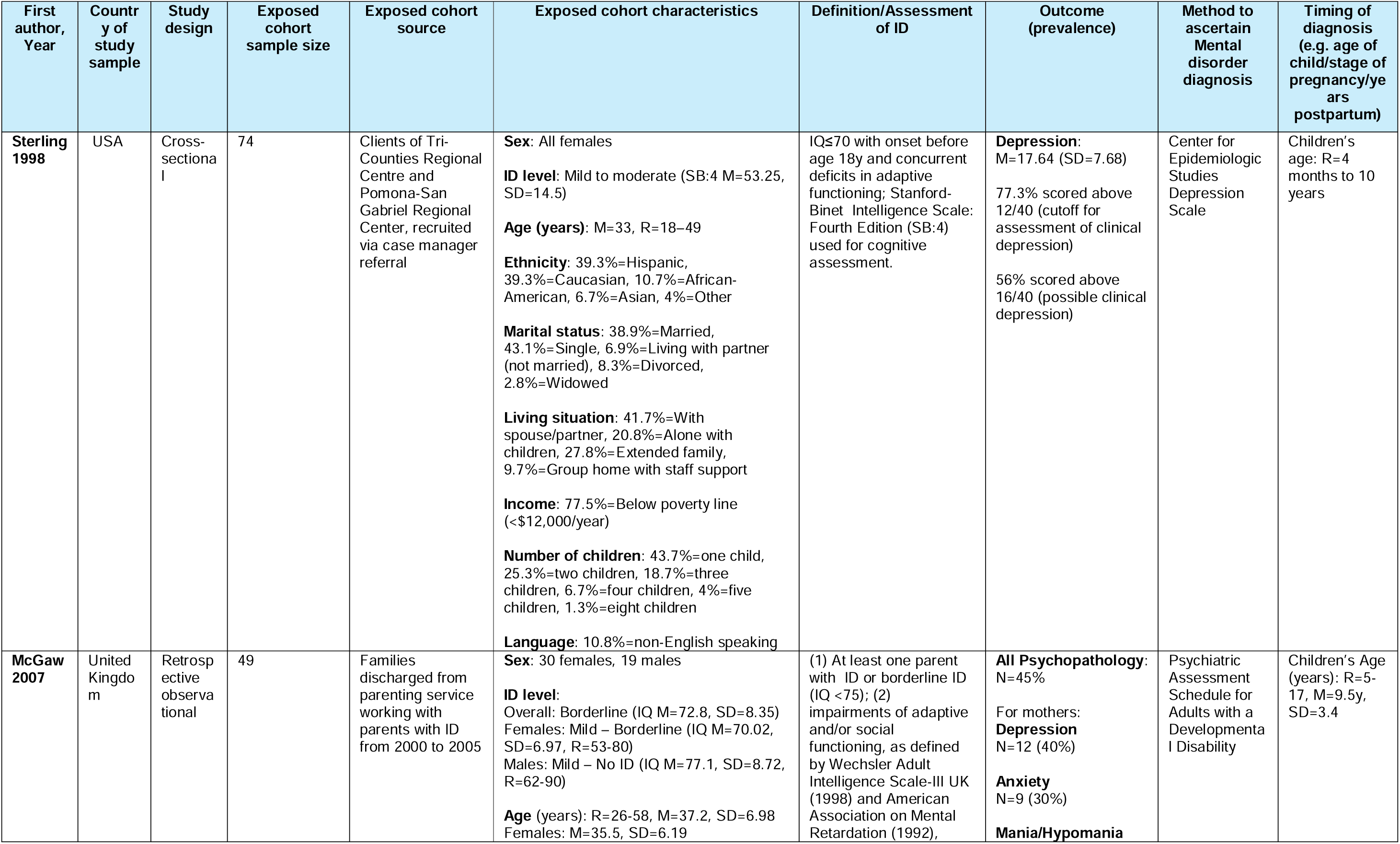
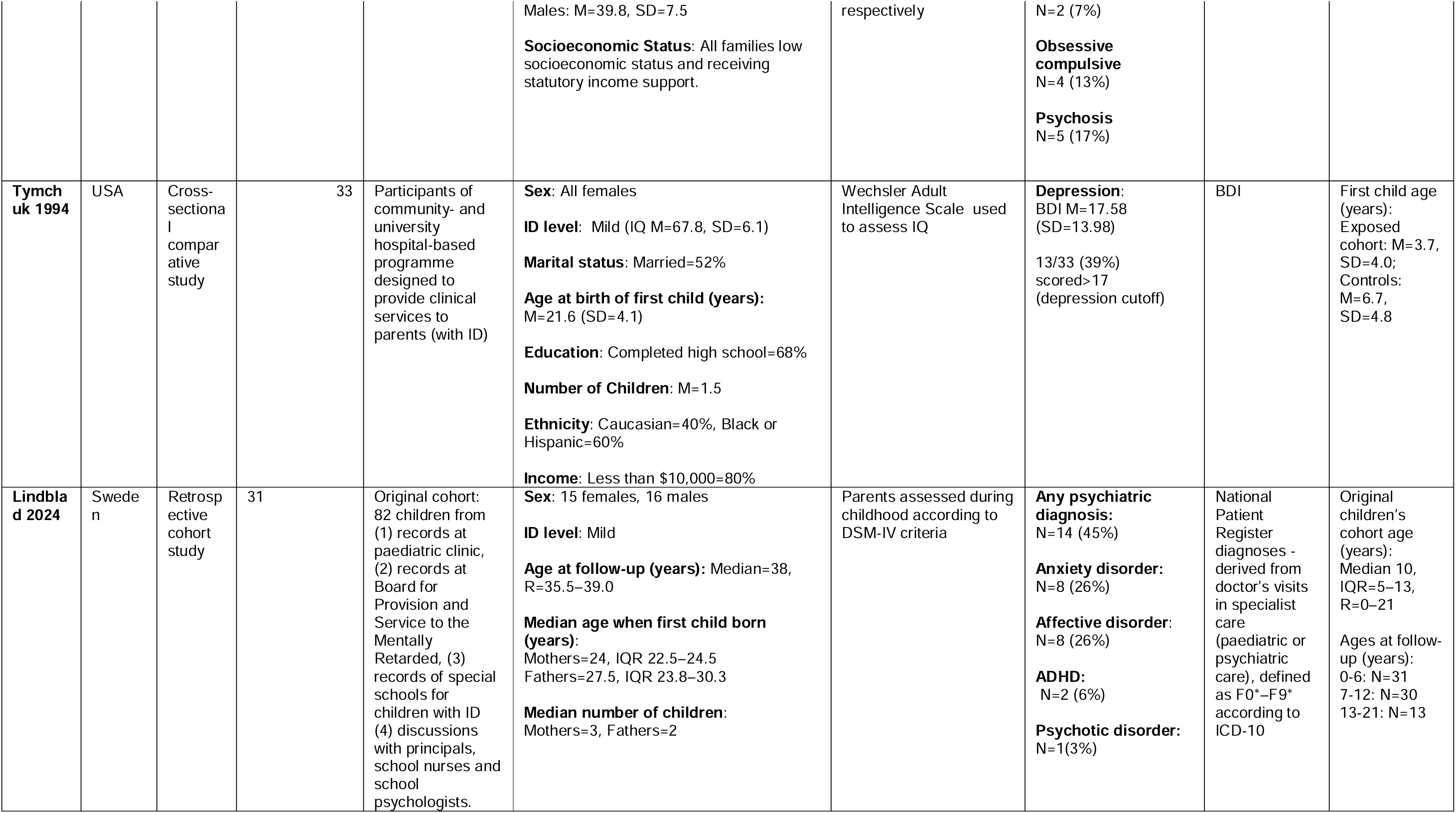
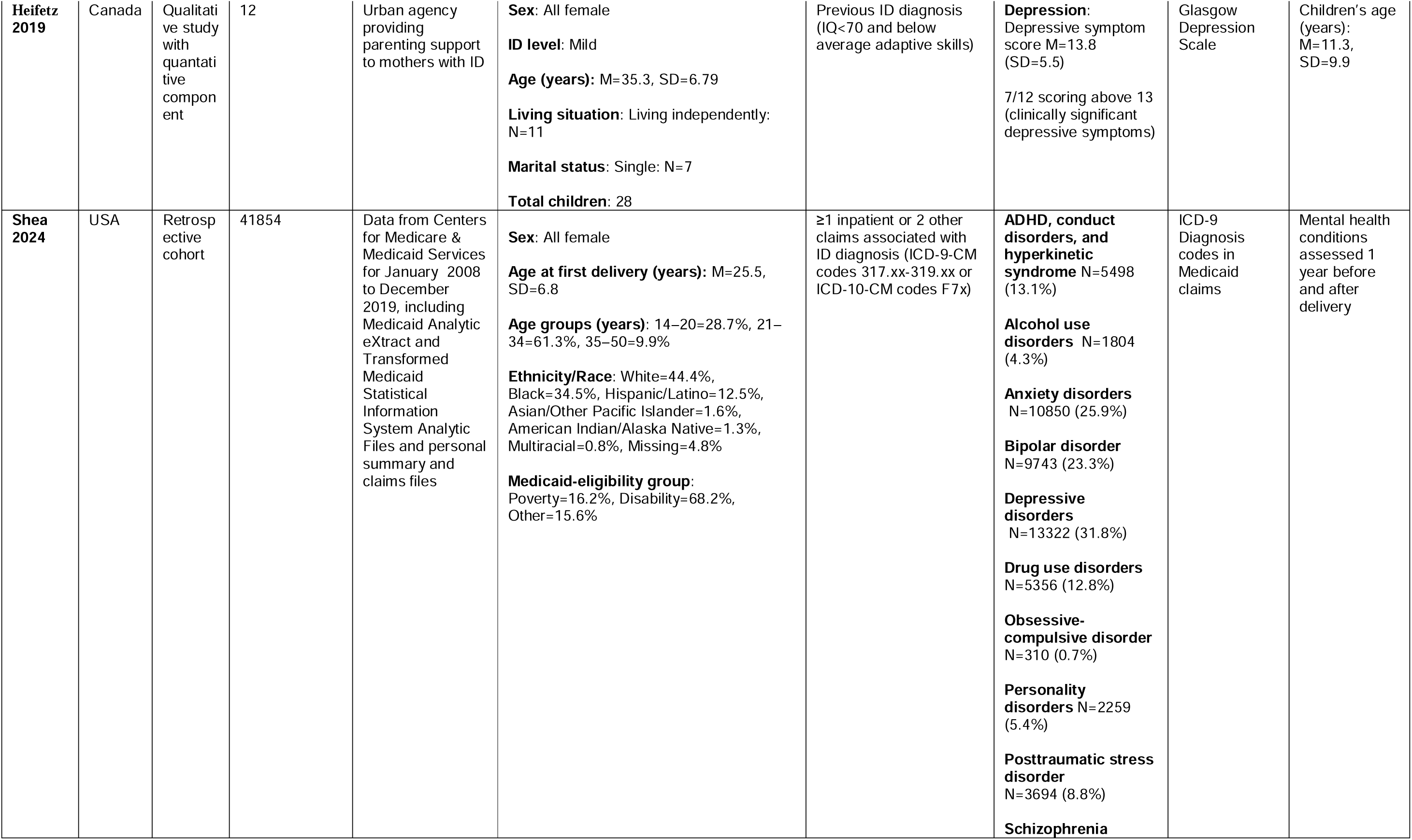
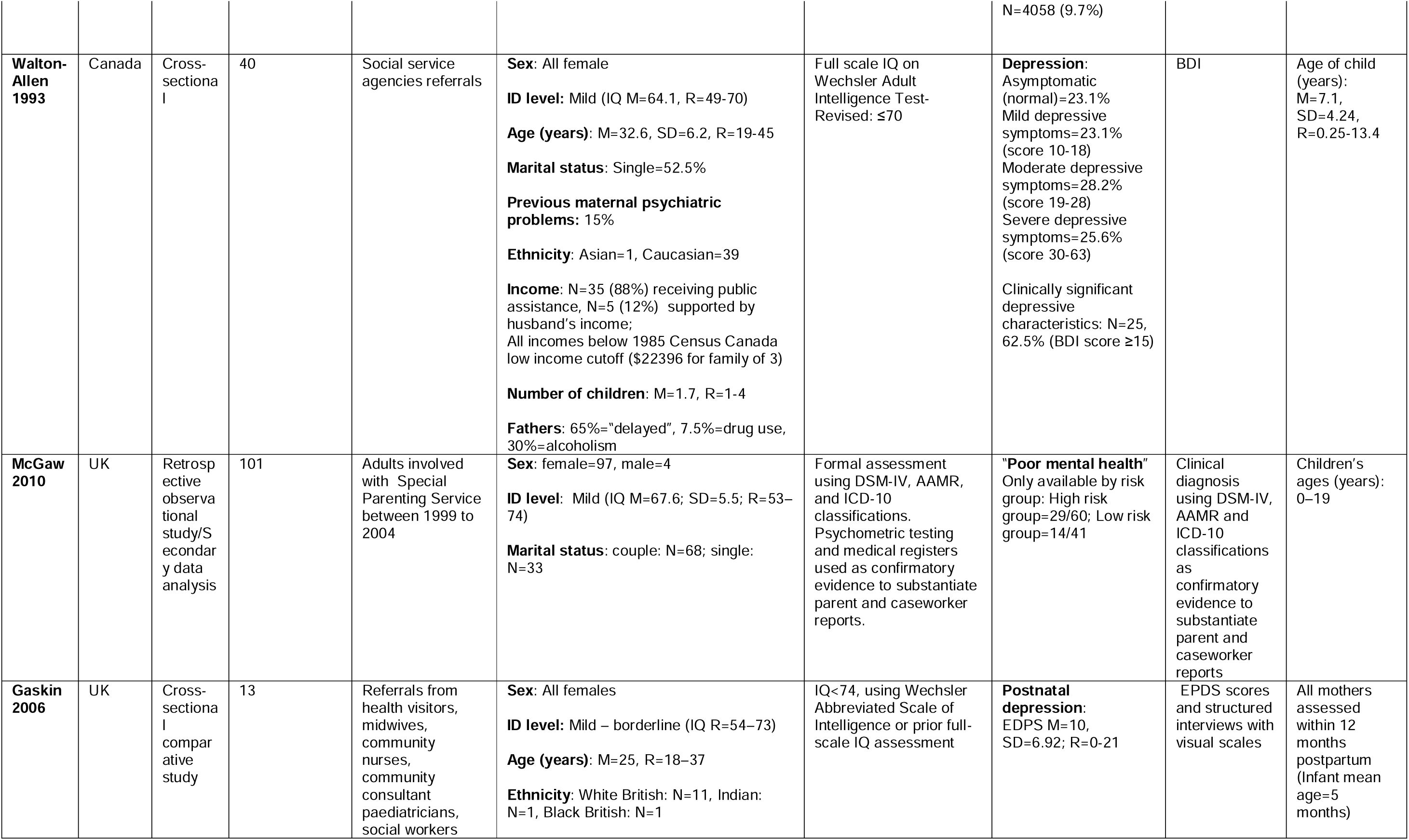
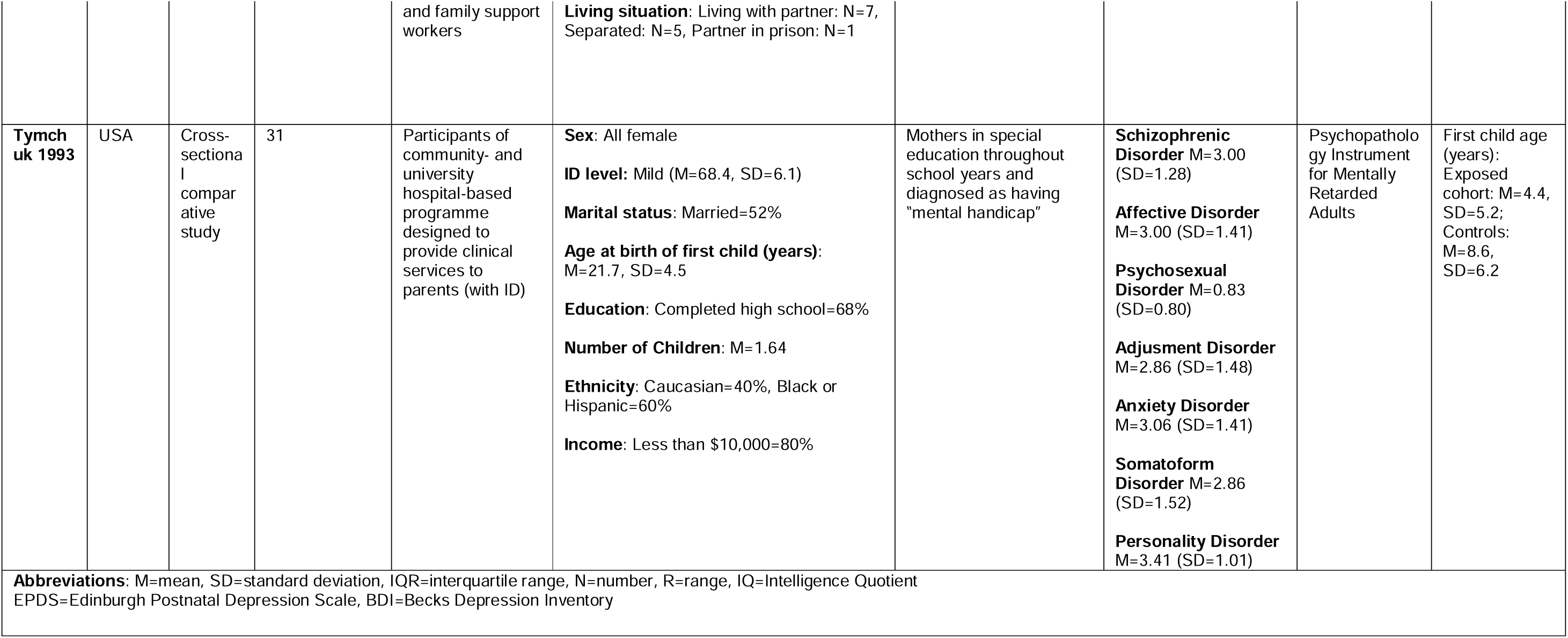
Characteristics of included studies. This table summarises data from the studies included in the review.

| First author, Year | Country of study sample | Study design | Exposed cohort sample size | Exposed cohort source | Exposed cohort characteristics | Definition/Assessment of ID | Outcome (prevalence) | Method to ascertain Mental disorder diagnosis | Timing of diagnosis (e.g. age of child/stage of pregnancy/years postpartum) |
| --- | --- | --- | --- | --- | --- | --- | --- | --- | --- |
| **Sterling 1998** | USA | Cross-sectional | 74 | Clients of Tri-Counties Regional Centre and Pomona-San Gabriel Regional Center, recruited via case manager referral | **Sex:** All females<br>**ID level:** Mild to moderate (SB:4 M=53.25, SD=14.5)<br>**Age (years):** M=33, R=18–49<br>**Ethnicity:** 39.3%=Hispanic, 39.3%=Caucasian, 10.7%=African-American, 6.7%=Asian, 4%=Other<br>**Marital status:** 38.9%=Married, 43.1%=Single, 6.9%=Living with partner (not married), 8.3%=Divorced, 2.8%=Widowed<br>**Living situation:** 41.7%=With spouse/partner, 20.8%=Alone with children, 27.8%=Extended family, 9.7%=Group home with staff support<br>**Income:** 77.5%=Below poverty line (<\$12,000/year)<br>**Number of children:** 43.7%=one child, 25.3%=two children, 18.7%=three children, 6.7%=four children, 4%=five children, 1.3%=eight children<br>**Language:** 10.8%=non-English speaking | IQ≤70 with onset before age 18y and concurrent deficits in adaptive functioning; Stanford-Binet Intelligence Scale: Fourth Edition (SB:4) used for cognitive assessment. | **Depression:** M=17.64 (SD=7.68)<br>77.3% scored above 12/40 (cutoff for assessment of clinical depression)<br>56% scored above 16/40 (possible clinical depression) | Center for Epidemiologic Studies Depression Scale | Children's age: R=4 months to 10 years |
| **McGaw 2007** | United Kingdom | Retrospective observational | 49 | Families discharged from parenting service working with parents with ID from 2000 to 2005 | **Sex:** 30 females, 19 males<br>**ID level:**<br>Overall: Borderline (IQ M=72.8, SD=8.35)<br>Females: Mild – Borderline (IQ M=70.02, SD=6.97, R=53-80)<br>Males: Mild – No ID (IQ M=77.1, SD=8.72, R=62-90)<br>**Age (years):** R=26-58, M=37.2, SD=6.98<br>Females: M=35.5, SD=6.19 | (1) At least one parent with ID or borderline ID (IQ <75); (2) impairments of adaptive and/or social functioning, as defined by Wechsler Adult Intelligence Scale-III UK (1998) and American Association on Mental Retardation (1992), | **All Psychopathology:** N=45%<br>For mothers:<br>**Depression** N=12 (40%)<br>**Anxiety** N=9 (30%)<br>**Mania/Hypomania** | Psychiatric Assessment Schedule for Adults with a Developmental Disability | Children's Age (years): R=5-17, M=9.5y, SD=3.4 |
|  |  |  |  |  | Males: M=39.8, SD=7.5<br>**Socioeconomic Status:** All families low socioeconomic status and receiving statutory income support. | respectively | N=2 (7%)<br>**Obsessive compulsive**<br>N=4 (13%)<br>**Psychosis**<br>N=5 (17%) |  |  |
| **Tymchuk 1994** | USA | Cross-sectional comparative study | 33 | Participants of community- and university hospital-based programme designed to provide clinical services to parents (with ID) | **Sex:** All females<br>**ID level:** Mild (IQ M=67.8, SD=6.1)<br>**Marital status:** Married=52%<br>**Age at birth of first child (years):** M=21.6 (SD=4.1)<br>**Education:** Completed high school=68%<br>**Number of Children:** M=1.5<br>**Ethnicity:** Caucasian=40%, Black or Hispanic=60%<br>**Income:** Less than \$10,000=80% | Wechsler Adult Intelligence Scale used to assess IQ | **Depression:**<br>BDI M=17.58 (SD=13.98)<br>13/33 (39%) scored > 17 (depression cutoff) | BDI | First child age (years):<br>Exposed cohort: M=3.7, SD=4.0;<br>Controls: M=6.7, SD=4.8 |
| **Lindblad 2024** | Sweden | Retrospective cohort study | 31 | Original cohort: 82 children from (1) records at paediatric clinic, (2) records at Board for Provision and Service to the Mentally Retarded, (3) records of special schools for children with ID (4) discussions with principals, school nurses and school psychologists. | **Sex:** 15 females, 16 males<br>**ID level:** Mild<br>**Age at follow-up (years):** Median=38, R=35.5–39.0<br>**Median age when first child born (years):**<br>Mothers=24, IQR 22.5–24.5<br>Fathers=27.5, IQR 23.8–30.3<br>**Median number of children:**<br>Mothers=3, Fathers=2 | Parents assessed during childhood according to DSM-IV criteria | **Any psychiatric diagnosis:**<br>N=14 (45%)<br>**Anxiety disorder:**<br>N=8 (26%)<br>**Affective disorder:**<br>N=8 (26%)<br>**ADHD:**<br>N=2 (6%)<br>**Psychotic disorder:**<br>N=1(3%) | National Patient Register diagnoses - derived from doctor's visits in specialist care (paediatric or psychiatric care), defined as F0*–F9* according to ICD-10 | Original children's cohort age (years):<br>Median 10, IQR=5–13, R=0–21<br>Ages at follow-up (years):<br>0-6: N=31<br>7-12: N=30<br>13-21: N=13 |
| **Heifetz 2019** | Canada | Qualitative study with quantitative component | 12 | Urban agency providing parenting support to mothers with ID | **Sex:** All female<br>**ID level:** Mild<br>**Age (years):** M=35.3, SD=6.79<br>**Living situation:** Living independently: N=11<br>**Marital status:** Single: N=7<br>**Total children:** 28 | Previous ID diagnosis (IQ<70 and below average adaptive skills) | **Depression:** Depressive symptom score M=13.8 (SD=5.5)<br>7/12 scoring above 13 (clinically significant depressive symptoms) | Glasgow Depression Scale | Children's age (years): M=11.3, SD=9.9 |
| **Shea 2024** | USA | Retrospective cohort | 41854 | Data from Centers for Medicare & Medicaid Services for January 2008 to December 2019, including Medicaid Analytic eXtract and Transformed Medicaid Statistical Information System Analytic Files and personal summary and claims files | **Sex:** All female<br>**Age at first delivery (years):** M=25.5, SD=6.8<br>**Age groups (years):** 14–20=28.7%, 21–34=61.3%, 35–50=9.9%<br>**Ethnicity/Race:** White=44.4%, Black=34.5%, Hispanic/Latino=12.5%, Asian/Other Pacific Islander=1.6%, American Indian/Alaska Native=1.3%, Multiracial=0.8%, Missing=4.8%<br>**Medicaid-eligibility group:** Poverty=16.2%, Disability=68.2%, Other=15.6% | ≥1 inpatient or 2 other claims associated with ID diagnosis (ICD-9-CM codes 317.xx-319.xx or ICD-10-CM codes F7x) | **ADHD, conduct disorders, and hyperkinetic syndrome** N=5498 (13.1%)<br>**Alcohol use disorders** N=1804 (4.3%)<br>**Anxiety disorders** N=10850 (25.9%)<br>**Bipolar disorder** N=9743 (23.3%)<br>**Depressive disorders** N=13322 (31.8%)<br>**Drug use disorders** N=5356 (12.8%)<br>**Obsessive-compulsive disorder** N=310 (0.7%)<br>**Personality disorders** N=2259 (5.4%)<br>**Posttraumatic stress disorder** N=3694 (8.8%)<br>**Schizophrenia** | ICD-9 Diagnosis codes in Medicaid claims | Mental health conditions assessed 1 year before and after delivery |
|  |  |  |  |  |  |  | N=4058 (9.7%) |  |  |
| **Walton-Allen 1993** | Canada | Cross-sectional | 40 | Social service agencies referrals | **Sex:** All female<br>**ID level:** Mild (IQ M=64.1, R=49-70)<br>**Age (years):** M=32.6, SD=6.2, R=19-45<br>**Marital status:** Single=52.5%<br>**Previous maternal psychiatric problems:** 15%<br>**Ethnicity:** Asian=1, Caucasian=39<br>**Income:** N=35 (88%) receiving public assistance, N=5 (12%) supported by husband's income; All incomes below 1985 Census Canada low income cutoff (\$22396 for family of 3)<br>**Number of children:** M=1.7, R=1-4<br>**Fathers:** 65%="delayed", 7.5%=drug use, 30%=alcoholism | Full scale IQ on Wechsler Adult Intelligence Test-Revised: ≤70 | **Depression:**<br>Asymptomatic (normal)=23.1%<br>Mild depressive symptoms=23.1% (score 10-18)<br>Moderate depressive symptoms=28.2% (score 19-28)<br>Severe depressive symptoms=25.6% (score 30-63)<br>Clinically significant depressive characteristics: N=25, 62.5% (BDI score ≥15) | BDI | Age of child (years): M=7.1, SD=4.24, R=0.25-13.4 |
| **McGaw 2010** | UK | Retrospective observational study/Secondary data analysis | 101 | Adults involved with Special Parenting Service between 1999 to 2004 | **Sex:** female=97, male=4<br>**ID level:** Mild (IQ M=67.6; SD=5.5; R=53-74)<br>**Marital status:** couple: N=68; single: N=33 | Formal assessment using DSM-IV, AAMR, and ICD-10 classifications. Psychometric testing and medical registers used as confirmatory evidence to substantiate parent and caseworker reports. | **"Poor mental health"**<br>Only available by risk group: High risk group=29/60; Low risk group=14/41 | Clinical diagnosis using DSM-IV, AAMR and ICD-10 classifications as confirmatory evidence to substantiate parent and caseworker reports | Children's ages (years): 0-19 |
| **Gaskin 2006** | UK | Cross-sectional comparative study | 13 | Referrals from health visitors, midwives, community nurses, community consultant paediatricians, social workers | **Sex:** All females<br>**ID level:** Mild – borderline (IQ R=54-73)<br>**Age (years):** M=25, R=18-37<br>**Ethnicity:** White British: N=11, Indian: N=1, Black British: N=1 | IQ<74, using Wechsler Abbreviated Scale of Intelligence or prior full-scale IQ assessment | **Postnatal depression:**<br>EDPS M=10, SD=6.92; R=0-21 | EPDS scores and structured interviews with visual scales | All mothers assessed within 12 months postpartum (Infant mean age=5 months) |
|  |  |  |  | and family support workers | **Living situation:** Living with partner: N=7, Separated: N=5, Partner in prison: N=1 |  |  |  |  |
| **Tymchuk 1993** | USA | Cross-sectional comparative study | 31 | Participants of community- and university hospital-based programme designed to provide clinical services to parents (with ID) | **Sex:** All female<br>**ID level:** Mild (M=68.4, SD=6.1)<br>**Marital status:** Married=52%<br>**Age at birth of first child (years):** M=21.7, SD=4.5<br>**Education:** Completed high school=68%<br>**Number of Children:** M=1.64<br>**Ethnicity:** Caucasian=40%, Black or Hispanic=60%<br>**Income:** Less than \$10,000=80% | Mothers in special education throughout school years and diagnosed as having "mental handicap" | **Schizophrenic Disorder** M=3.00 (SD=1.28)<br>**Affective Disorder** M=3.00 (SD=1.41)<br>**Psychosexual Disorder** M=0.83 (SD=0.80)<br>**Adjustment Disorder** M=2.86 (SD=1.48)<br>**Anxiety Disorder** M=3.06 (SD=1.41)<br>**Somatoform Disorder** M=2.86 (SD=1.52)<br>**Personality Disorder** M=3.41 (SD=1.01) | Psychopathology Instrument for Mentally Retarded Adults | First child age (years): Exposed cohort: M=4.4, SD=5.2; Controls: M=8.6, SD=6.2 |
| **Abbreviations:** M=mean, SD=standard deviation, IQR=interquartile range, N=number, R=range, IQ=Intelligence Quotient<br>EPDS=Edinburgh Postnatal Depression Scale, BDI=Becks Depression Inventory |  |  |  |  |  |  |  |  |  |

The studies included a total of 42,207 individuals (excluding any overlapping participants) aged between 18 and 58 years. ID had been diagnosed based on DSM-IV criteria (Lindblad et al., 2024; McGaw et al., 2010; Sterling, 1998); ICD-9 or ICD-10 criteria (McGaw et al., 2010; Shea et al., 2024); American Association on Mental Retardation 1992 Manual (McGaw et al., 2010; McGaw et al., 2007); American Association on Intellectual and Developmental Disabilities Guidance 2011 (Heifetz et al., 2019); and cognitive testing using Wechsler Adult Intelligence Scale or Wechsler Abbreviated Scale of Intelligence (Gaskin & James, 2006; Tymchuk, 1993; Tymchuk, 1994; Walton-Allen, 1993) - The method of diagnosing ID focusing on cognitive testing is in line with ICD-10 and DSM-III, which were in use when these studies were published. There were no studies identified including participants with genetic syndromes.

The majority of studies included mothers only. Three studies included fathers: Lindblad et al. (2024) included 16 fathers and 15 mothers; McGaw et al. (2010) included four fathers and 97 mothers; McGaw et al. (2007) included 19 couples in which at least one had ID as well as an additional 11 females. In McGaw et al. (2007) it was unclear how many of the fathers had ID - The mean IQ presented for males was above what is expected for an ID diagnoses (70 ± 5 points on standardised tests), whilst female mean IQ was compatible with ID diagnosis (Males: M=77.1, SD=8.72, range=62-90; Females: M=70.02, SD=6.97, range=53-80). Therefore, the results for females rather than for males were extracted from this study.

The studies identified provided sample prevalences for psychiatric disorders overall (Lindblad et al., 2024; McGaw et al., 2010; McGaw et al., 2007), as well as specific prevalences for depression (Heifetz et al., 2019; McGaw et al., 2007; Shea et al., 2024; Sterling, 1998; Tymchuk, 1994; Walton-Allen, 1993), anxiety (Lindblad et al., 2024; McGaw et al., 2007; Shea et al., 2024), psychosis/psychotic disorder/schizophrenia (Lindblad et al., 2024; McGaw et al., 2007; Shea et al., 2024), mania/bipolar affective disorder (McGaw et al., 2007; Shea et al., 2024), Obsessive Compulsive Disorder (OCD) (McGaw et al., 2007; Shea et al., 2024), Post Traumatic Stress Disorder (PTSD) (Shea et al., 2024) and alcohol/drug use disorders (Shea et al., 2024). The majority of studies used screening tools to assess the presence of mental disorders apart from three which used clinical diagnoses (Lindblad et al., 2024; McGaw et al., 2010; Shea et al., 2024). The prevalence of a mental disorder in the sample was given in nine out of eleven of the studies. In the other two (Gaskin & James, 2006; Tymchuk, 1993), mean scores on screening scales were given without details of the clinical cut-off for a probable diagnosis. Therefore, these studies were not included in the meta-analysis but included in a narrative synthesis.

None of the identified studies mentioned exclusively analysing diagnoses made during pregnancy, although mental health conditions were assessed one year before and after delivery in Shea et al. (2024). The only study to administer screening tools to mothers within 12 months post-partum was Gaskin and James (2006). The remaining studies used screening tools and diagnoses with women with children aged between 0-21 years.

### 3.3. Risk of Bias

There were concerns regarding the risk of bias for all studies, with over half assessed as “high risk of bias” (Gaskin & James, 2006; Heifetz et al., 2019; McGaw et al., 2007; Sterling, 1998; Tymchuk, 1993; Walton-Allen, 1993) and four as “moderate risk of bias” (Lindblad et al., 2024; McGaw et al., 2010; Shea et al., 2024; Tymchuk, 1994). In general studies were at high risk of bias due to their restrictive sampling approaches, small non-random samples and missing numbers for participants that did not respond/consent to take part. Detailed breakdown of MMAT results and judgement justifications are available in supplementary materials (S3).

### 3.4. Prevalence of Mental Disorders in Parents with ID

#### 3.4.1. Results of Meta-Analyses

For parents with ID, the prevalence of any psychiatric disorder was 46% (95% CI 38–53) with I^2^ 0.0%, p=0.391 (**Figure 2**). Depression showed significant heterogeneity across eras (I^2^=86.7%, p<0.001); however in pre-2000 studies, pooled prevalence of depression was 53% (95% CI: 41–65%) with moderate heterogeneity (I²=52.8%, p=0.120) and post-2000 was 38% (95% CI: 26–50%), with moderate heterogeneity (I²=53.5%, p=0.117) (**Figure 3**). Pooled anxiety was 26% (95% CI:26– 26%) with no variation attributable to heterogeneity (I^2^=0.0%, p=0.888). For psychosis, the pooled prevalence was 9% (95% CI:3–14%) with moderate variation attributable to heterogeneity (I^2^=61.6%, p=0.074). Data for bipolar disorder, OCD, PTSD, and substance use were sparse. (Forest plots for depression, anxiety, and psychosis available in supplementary material S4a-c.)

**Figure 2:**
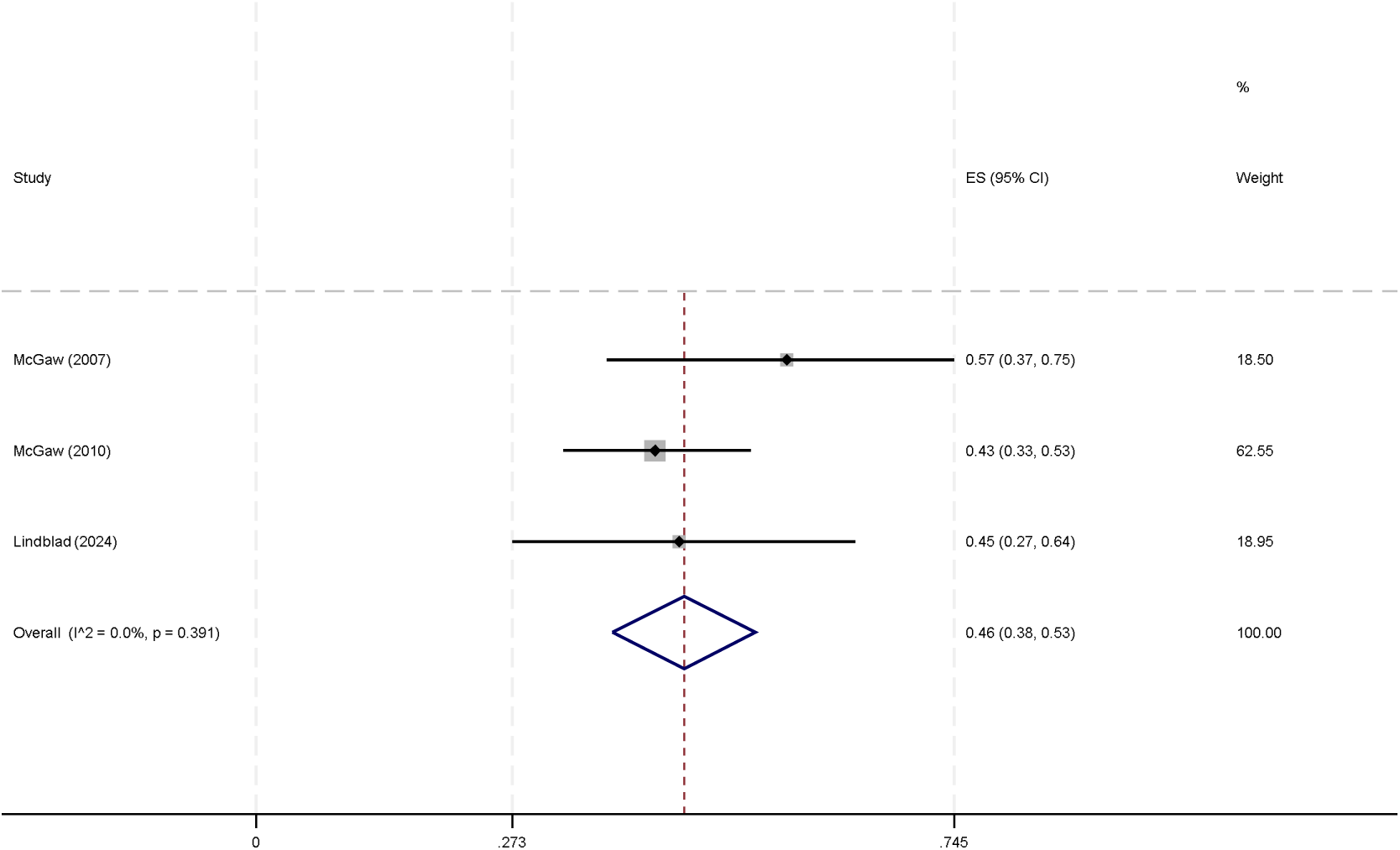
Forest plot showing studies providing prevalence figures for any psychiatric disorder in parents with ID. Three studies provided the sample prevalences for psychiatric disorders overall in parents with ID. The pooled prevalence was 46% (95% CI: 38-53%) with I^2^ 0.0%, p=0.391.

**Figure 3:**
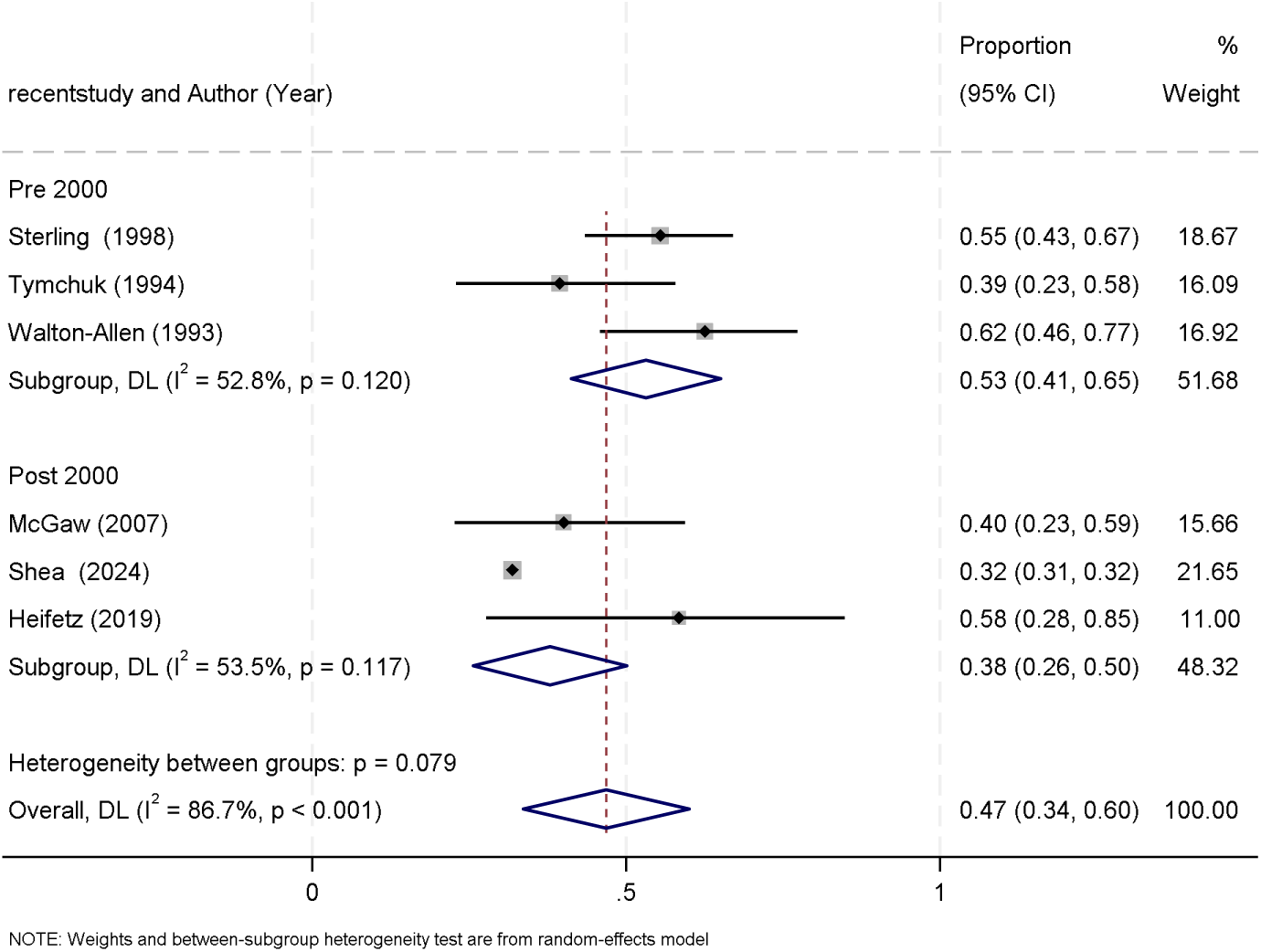
Forest plot showing studies providing prevalence figures for depression pre and post-2000 in parents with ID. Studies reporting the prevalence of depression conducted after 2000 yielded a prevalence of 38% (95% CI: 26–50%), with moderate heterogeneity (I² = 53.5%, p = 0.117). In contrast, studies conducted prior to 2000 demonstrated a higher prevalence of 53% (95% CI: 41–65%), also with moderate heterogeneity (I² = 52.8%, p = 0.120).

#### 3.4.2. Screening Tool Findings

Across smaller studies using Centre Epidemiologic Studies Depression Scale (CES-D), Glasgow Depression Scale (GDS) and Beck Depression Inventory (BDI), many mothers scored above thresholds for possible clinical depression (Heifetz et al., 2019; Sterling, 1998; Tymchuk, 1994), whilst lack of validated cutl7loffs in other studies (Gaskin & James, 2006; Tymchuk, 1993) limited their interpretability.

#### 3.4.3. Comparative Studies

A large US administrative cohort found substantially higher adjusted odds ratios among birthing people with ID versus those without IDD: anxiety disorders 2.49 (2.40-2.58); depressive disorders 3.01 (2.91-3.10); bipolar disorder 3.79 (3.64-3.95); PTSD 3.38 (3.17-3.60); OCD 2.38 (1.95-2.91); schizophrenia 7.93 (7.30-8.62); alcohol use disorders 2.02 (1.87-2.18); and drug use disorders 1.42 (1.36-1.49) (Shea et al., 2024). Two small 1990s studies (Tymchuk, 1993; Tymchuk, 1994) reported significantly higher scores in mothers with ID versus mothers without ID on screening for schizophrenic disorder, affective disorder, adjustment disorder, anxiety disorder and depression. In contrast, a small Swedish register study (Lindblad et al., 2024) comparing parents with ID to nonl7lparents with ID found no significant differences. These findings are summarised in **Table 2**.

**Table 2:**
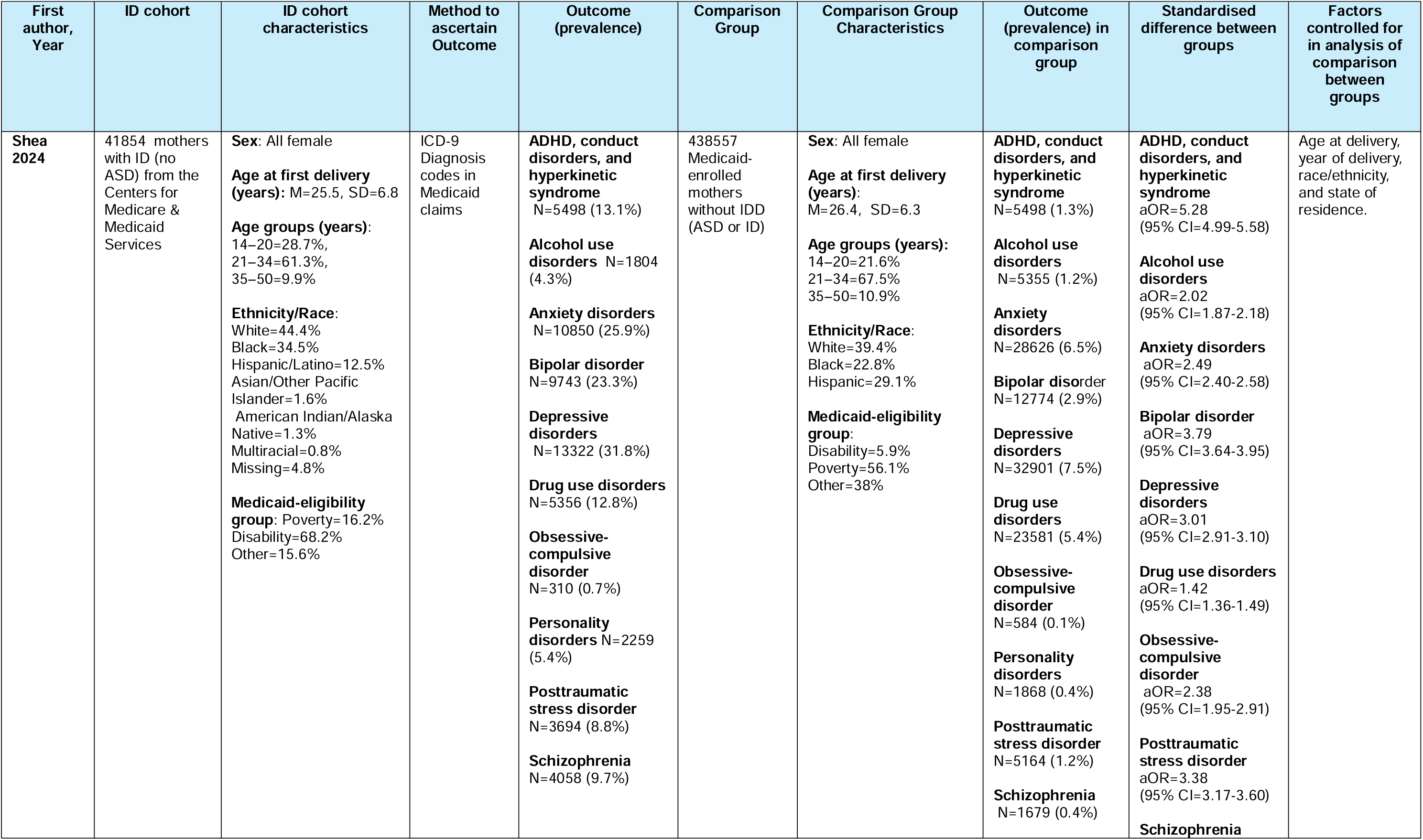
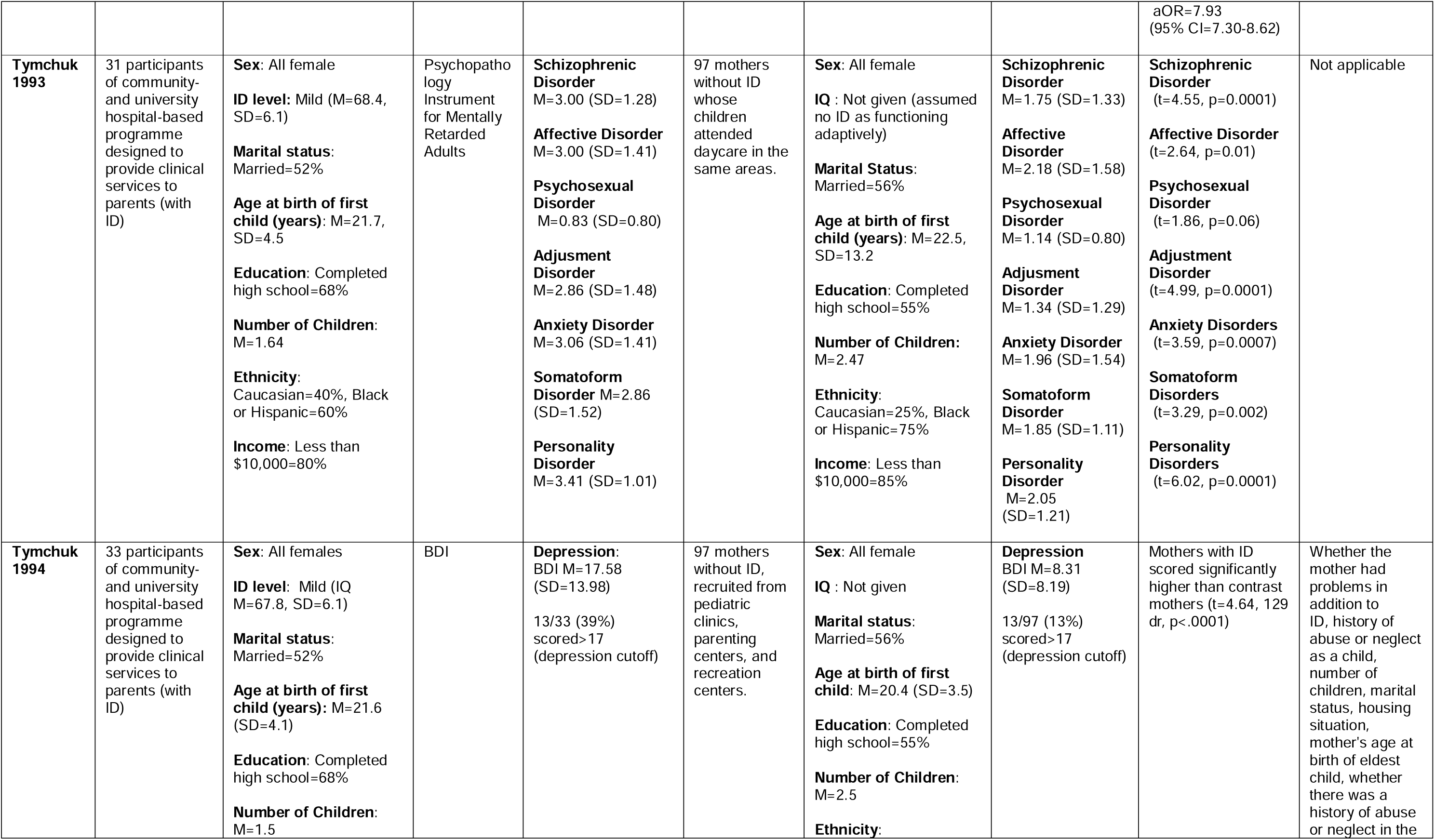
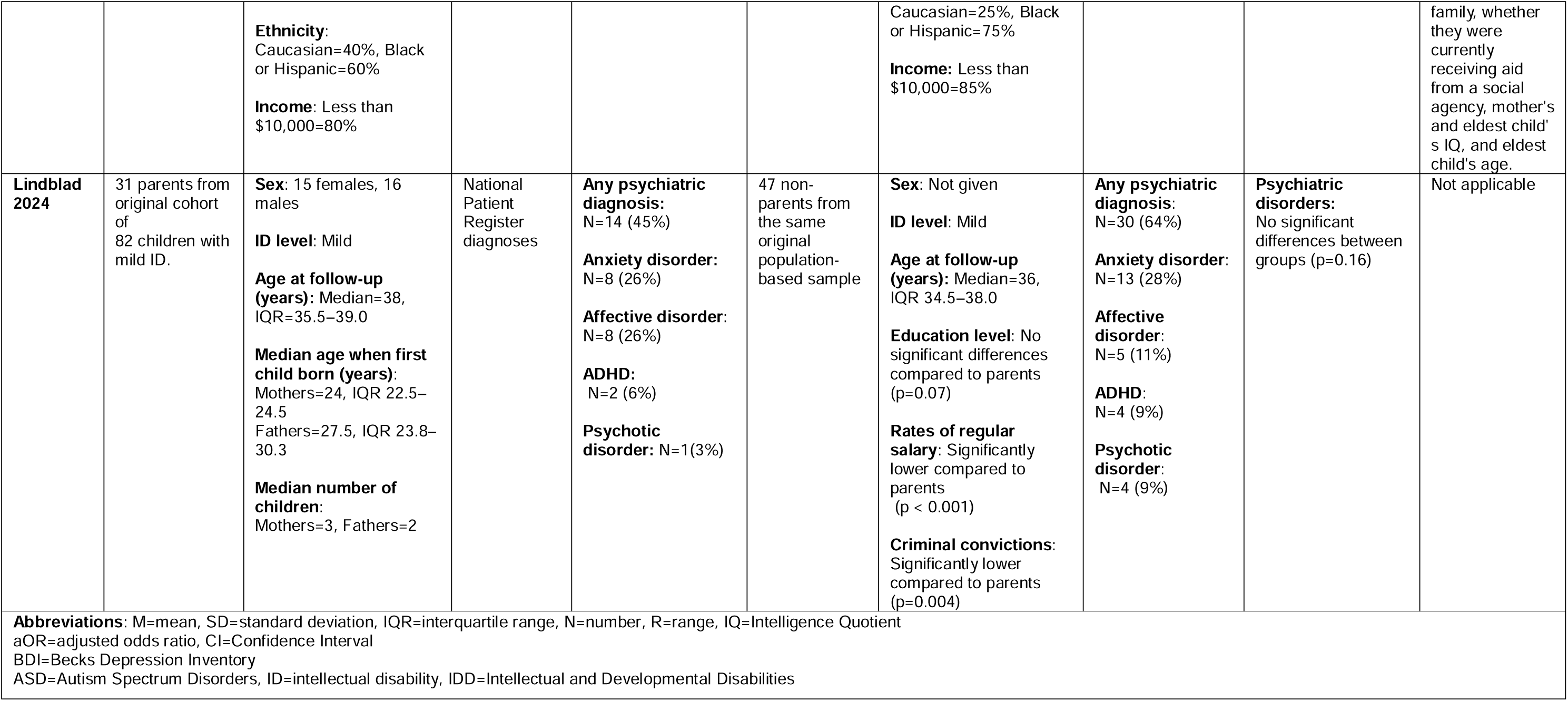
Results from comparison studies. Comparisons of rates of mental disorders in mothers with ID with those without ID or non-parents with ID was provided in four of the studies summarised in the table.

| First author, Year | ID cohort | ID cohort characteristics | Method to ascertain Outcome | Outcome (prevalence) | Comparison Group | Comparison Group Characteristics | Outcome (prevalence) in comparison group | Standardised difference between groups | Factors controlled for in analysis of comparison between groups |
| --- | --- | --- | --- | --- | --- | --- | --- | --- | --- |
| Shea 2024 | 41854 mothers with ID (no ASD) from the Centers for Medicare & Medicaid Services | **Sex:** All female<br>**Age at first delivery (years):** M=25.5, SD=6.8<br>**Age groups (years):**<br>14–20=28.7%,<br>21–34=61.3%,<br>35–50=9.9%<br>**Ethnicity/Race:**<br>White=44.4%<br>Black=34.5%<br>Hispanic/Latino=12.5%<br>Asian/Other Pacific Islander=1.6%<br>American Indian/Alaska Native=1.3%<br>Multiracial=0.8%<br>Missing=4.8%<br>**Medicaid-eligibility group:** Poverty=16.2%<br>Disability=68.2%<br>Other=15.6% | ICD-9 Diagnosis codes in Medicaid claims | **ADHD, conduct disorders, and hyperkinetic syndrome**<br>N=5498 (13.1%)<br>**Alcohol use disorders** N=1804 (4.3%)<br>**Anxiety disorders**<br>N=10850 (25.9%)<br>**Bipolar disorder**<br>N=9743 (23.3%)<br>**Depressive disorders**<br>N=13322 (31.8%)<br>**Drug use disorders**<br>N=5356 (12.8%)<br>**Obsessive-compulsive disorder**<br>N=310 (0.7%)<br>**Personality disorders** N=2259 (5.4%)<br>**Posttraumatic stress disorder**<br>N=3694 (8.8%)<br>**Schizophrenia**<br>N=4058 (9.7%) | 438557 Medicaid-enrolled mothers without IDD (ASD or ID) | **Sex:** All female<br>**Age at first delivery (years):**<br>M=26.4, SD=6.3<br>**Age groups (years):**<br>14–20=21.6%<br>21–34=67.5%<br>35–50=10.9%<br>**Ethnicity/Race:**<br>White=39.4%<br>Black=22.8%<br>Hispanic=29.1%<br>**Medicaid-eligibility group:**<br>Disability=5.9%<br>Poverty=56.1%<br>Other=38% | **ADHD, conduct disorders, and hyperkinetic syndrome**<br>N=5498 (1.3%)<br>**Alcohol use disorders**<br>N=5355 (1.2%)<br>**Anxiety disorders**<br>N=28626 (6.5%)<br>**Bipolar disorder**<br>N=12774 (2.9%)<br>**Depressive disorders**<br>N=32901 (7.5%)<br>**Drug use disorders**<br>N=23581 (5.4%)<br>**Obsessive-compulsive disorder**<br>N=584 (0.1%)<br>**Personality disorders**<br>N=1868 (0.4%)<br>**Posttraumatic stress disorder**<br>N=5164 (1.2%)<br>**Schizophrenia**<br>N=1679 (0.4%) | **ADHD, conduct disorders, and hyperkinetic syndrome**<br>aOR=5.28<br>(95% CI=4.99-5.58)<br>**Alcohol use disorders**<br>aOR=2.02<br>(95% CI=1.87-2.18)<br>**Anxiety disorders**<br>aOR=2.49<br>(95% CI=2.40-2.58)<br>**Bipolar disorder**<br>aOR=3.79<br>(95% CI=3.64-3.95)<br>**Depressive disorders**<br>aOR=3.01<br>(95% CI=2.91-3.10)<br>**Drug use disorders**<br>aOR=1.42<br>(95% CI=1.36-1.49)<br>**Obsessive-compulsive disorder**<br>aOR=2.38<br>(95% CI=1.95-2.91)<br>**Posttraumatic stress disorder**<br>aOR=3.38<br>(95% CI=3.17-3.60)<br>**Schizophrenia** aOR=7.93<br>(95% CI=7.30-8.62) | Age at delivery, year of delivery, race/ethnicity, and state of residence. |
| **Tymchuk 1993** | 31 participants of community- and university hospital-based programme designed to provide clinical services to parents (with ID) | **Sex:** All female<br>**ID level:** Mild (M=68.4, SD=6.1)<br>**Marital status:** Married=52%<br>**Age at birth of first child (years):** M=21.7, SD=4.5<br>**Education:** Completed high school=68%<br>**Number of Children:** M=1.64<br>**Ethnicity:** Caucasian=40%, Black or Hispanic=60%<br>**Income:** Less than \$10,000=80% | Psychopathology Instrument for Mentally Retarded Adults | **Schizophrenic Disorder** M=3.00 (SD=1.28)<br>**Affective Disorder** M=3.00 (SD=1.41)<br>**Psychosexual Disorder** M=0.83 (SD=0.80)<br>**Adjustment Disorder** M=2.86 (SD=1.48)<br>**Anxiety Disorder** M=3.06 (SD=1.41)<br>**Somatoform Disorder** M=2.86 (SD=1.52)<br>**Personality Disorder** M=3.41 (SD=1.01) | 97 mothers without ID whose children attended daycare in the same areas. | **Sex:** All female<br>**IQ :** Not given (assumed no ID as functioning adaptively)<br>**Marital Status:** Married=56%<br>**Age at birth of first child (years):** M=22.5, SD=13.2<br>**Education:** Completed high school=55%<br>**Number of Children:** M=2.47<br>**Ethnicity:** Caucasian=25%, Black or Hispanic=75%<br>**Income:** Less than \$10,000=85% | **Schizophrenic Disorder** M=1.75 (SD=1.33)<br>**Affective Disorder** M=2.18 (SD=1.58)<br>**Psychosexual Disorder** M=1.14 (SD=0.80)<br>**Adjustment Disorder** M=1.34 (SD=1.29)<br>**Anxiety Disorder** M=1.96 (SD=1.54)<br>**Somatoform Disorder** M=1.85 (SD=1.11)<br>**Personality Disorder** M=2.05 (SD=1.21) | **Schizophrenic Disorder** (t=4.55, p=0.0001)<br>**Affective Disorder** (t=2.64, p=0.01)<br>**Psychosexual Disorder** (t=1.86, p=0.06)<br>**Adjustment Disorder** (t=4.99, p=0.0001)<br>**Anxiety Disorders** (t=3.59, p=0.0007)<br>**Somatoform Disorders** (t=3.29, p=0.002)<br>**Personality Disorders** (t=6.02, p=0.0001) | Not applicable |
| **Tymchuk 1994** | 33 participants of community- and university hospital-based programme designed to provide clinical services to parents (with ID) | **Sex:** All females<br>**ID level:** Mild (IQ M=67.8, SD=6.1)<br>**Marital status:** Married=52%<br>**Age at birth of first child (years):** M=21.6 (SD=4.1)<br>**Education:** Completed high school=68%<br>**Number of Children:** M=1.5 | BDI | **Depression:** BDI M=17.58 (SD=13.98)<br>13/33 (39%) scored > 17 (depression cutoff) | 97 mothers without ID, recruited from pediatric clinics, parenting centers, and recreation centers. | **Sex:** All female<br>**IQ :** Not given<br>**Marital status:** Married=56%<br>**Age at birth of first child:** M=20.4 (SD=3.5)<br>**Education:** Completed high school=55%<br>**Number of Children:** M=2.5<br>**Ethnicity:** | **Depression** BDI M=8.31 (SD=8.19)<br>13/97 (13%) scored > 17 (depression cutoff) | Mothers with ID scored significantly higher than contrast mothers (t=4.64, 129 df, p<.0001) | Whether the mother had problems in addition to ID, history of abuse or neglect as a child, number of children, marital status, housing situation, mother's age at birth of eldest child, whether there was a history of abuse or neglect in the |
| | | **Ethnicity:**<br>Caucasian=40%, Black or Hispanic=60%<br>**Income:** Less than \$10,000=80% | | | | Caucasian=25%, Black or Hispanic=75%<br>**Income:** Less than \$10,000=85% | | | family, whether they were currently receiving aid from a social agency, mother's and eldest child's IQ, and eldest child's age. |
| **Lindblad 2024** | 31 parents from original cohort of 82 children with mild ID. | **Sex:** 15 females, 16 males<br>**ID level:** Mild<br>**Age at follow-up (years):** Median=38, IQR=35.5–39.0<br>**Median age when first child born (years):** Mothers=24, IQR 22.5–24.5<br>Fathers=27.5, IQR 23.8–30.3<br>**Median number of children:** Mothers=3, Fathers=2 | National Patient Register diagnoses | **Any psychiatric diagnosis:** N=14 (45%)<br>**Anxiety disorder:** N=8 (26%)<br>**Affective disorder:** N=8 (26%)<br>**ADHD:** N=2 (6%)<br>**Psychotic disorder:** N=1(3%) | 47 non-parents from the same original population-based sample | **Sex:** Not given<br>**ID level:** Mild<br>**Age at follow-up (years):** Median=36, IQR 34.5–38.0<br>**Education level:** No significant differences compared to parents (p=0.07)<br>**Rates of regular salary:** Significantly lower compared to parents (p < 0.001)<br>**Criminal convictions:** Significantly lower compared to parents (p=0.004) | **Any psychiatric diagnosis:** N=30 (64%)<br>**Anxiety disorder:** N=13 (28%)<br>**Affective disorder:** N=5 (11%)<br>**ADHD:** N=4 (9%)<br>**Psychotic disorder:** N=4 (9%) | **Psychiatric disorders:** No significant differences between groups (p=0.16) | Not applicable |
| **Abbreviations:** M=mean, SD=standard deviation, IQR=interquartile range, N=number, R=range, IQ=Intelligence Quotient<br>aOR=adjusted odds ratio, CI=Confidence Interval<br>BDI=Becks Depression Inventory<br>ASD=Autism Spectrum Disorders, ID=intellectual disability, IDD=Intellectual and Developmental Disabilities |  |  |  |  |  |  |  |  |  |

#### 3.4.4. Comparisons with Wider Literature

Across the studies of parents with ID, 46% were found to have psychiatric disorders, in comparison to rates of psychiatric disorders in those with mild ID (“mild” being the level of ID most commonly impacting the ID parent sample) of 29.1% (Mazza et al., 2020), compared to 16% of the adult general population (Cooper et al., 2007). 38% of mothers with ID experience depression, compared to 9.4% of people with mild ID (Maïano et al., 2018), up to 22.0% of females with ID (Carey et al., 2017), and 5.7% of the general adult population (World Health Organization, 2025). 26% of parents with ID experienced anxiety, compared to 5.5% across the lifespan in people with ID (Maïano et al., 2018; Mazza et al., 2020; Oeseburg et al., 2011), and 4.05% of the general population (Javaid et al., 2023). 9% of parents with ID experienced psychosis, compared to 5.6% in people with mild ID (Aman et al., 2016), and 0.3%-0.7% in the general population (Rahman & Lauriello, 2016).

McGaw et al. (2007) reported 7% mania/hypomania in mothers with ID and Shea et al. (2024) reported 23.3% bipolar disorder in mothers with ID, compared to 2.9% in mothers without ID. These estimates differ, likely due to the different study designs, populations included, and methods used. In comparison the lifetime prevalence of bipolar disorder (types one and two combined) is 3.6%-6.2% in autistic adults with ID (Varcin et al., 2022), and 2.6% in the general population (Clemente et al., 2015).

McGaw et al. (2007) and Shea et al. (2024) also presented different results for OCD in mothers with ID (13% and 0.7% respectively), compared to 0.1% of mothers without ID. In comparison, the point prevalence of OCD in females with mild ID has been reported up to 1.5% (Cooper et al., 2007).

The one study to report on PTSD and substance use disorders in mothers with ID reported 8.8% PTSD in mothers with ID compared to 1.2% in mothers without ID (Shea et al., 2024). In comparison, Daveney et al. (2019) reported a higher prevalence of PTSD in adults with ID of 10.0% (95% CI: 0.4%-19.5%). Shea et al. (2024) also reported 4.3% and 12.8% of mothers with ID with alcohol and drug use disorders respectively, compared to 1.2% and 5.4% of mothers without ID. In comparison, a lower point prevalence of 1.0% alcohol/substance use disorder in females with mild ID has been reported (Cooper et al., 2007).

## 4. DISCUSSION

### 4.1. Key Findings

Across studies included in this review, approximately half of parents with ID— including mothers and fathers—were found to experience a psychiatric disorder. Depression was the most frequently studied outcome, with 38% of mothers experiencing depressive symptoms in studies conducted post-2000. Anxiety and psychosis were less common but still affected a substantial proportion of parents, with prevalence rates of 26% and 9%, respectively. These findings indicate a considerable mental health burden within this population.

This review shows that parents with ID have a higher prevalence of mental disorders than both the general population and individuals with ID who are not parents. This aligns with previous research; for example, a review of mental health outcomes across the reproductive life course among women with disabilities also found heightened risks of poor mental health among those with IDD (Deierlein et al., 2024). While that review employed a broader definition of ID—including self-report and other assessment methods—and restricted inclusion to studies with comparison groups, its conclusions are consistent with the present findings, further highlighting the disproportionate vulnerability of parents with ID to adverse mental health outcomes.

### 4.2. Strengths and Limitations

Definitions of ID varied across studies, reflecting shifts in diagnostic practice, and requiring a formal diagnosis may have excluded undiagnosed parents. Outcomes were also assessed inconsistently between studies (diagnostic codes vs. screening tools), and many studies were small or high risk of bias. Nonetheless, this is the first systematic review to offer pooled estimates to demonstrate elevated risks of mental disorders among parents with ID, thus advancing our understanding of mental health outcomes in this population and identifying key gaps in the literature that require further investigation.

### 4.3. Evidence Gaps and Future Research

There is an overall lack of literature examining mental disorders in parents with ID, with initial identification of nearly 13,000 records yielding only ten records suitable for the review. This included studies that were rated as high risk of bias, two doctoral theses and two studies that did not provide data in a form that could be meta-analysed. This sparsity of literature overall meant we were unable to perform subgroup analyses. Larger studies are required using population-based data to ensure a greater representativeness of findings.

Only two studies gave data for mania/bipolar affective disorder and OCD (McGaw et al., 2007; Shea et al., 2024) and one study gave data for PTSD and alcohol/drug use disorders (Shea et al., 2024). Given the conflicting results between these two studies, concerns regarding risk of bias and significant heterogeneity, more studies are required to aid accurate interpretation of the prevalence of these disorders in this population.

The review highlighted a particularly concerning gap in the knowledge base on mental disorders in mothers with ID within the context of pregnancy and post-partum, with the majority of studies including a mixture of women with children from ages 0-21 years. Whilst the postpartum period is traditionally defined as the first six weeks following childbirth (Lopez-Gonzalez & Kopparapu, 2022), many contemporary guidelines recognize that maternal recovery extends beyond this period, hence specialist services may stay involved for up to one year postpartum (Saldanha et al., 2023). Furthermore, no studies focused on mental disorders in women with ID who had experienced pregnancies that did not result in live births. It is important that studies focus on these periods where women are particularly vulnerable.

The review also indicates a lack of literature pertaining to mental disorders in fathers with ID. Qualitative studies including fathers with ID have highlighted that fathers also face mental health challenges which can impact their abilities to parent and support their partners (Symonds et al., 2021). Therefore, quantitative studies should include fathers, in order for services to learn how to provide them with necessary tailored support.

There was a gap in literature comparing outcomes for parents with ID compared to non-parents with ID. Only Lindblad (2024) compared parents with ID to non-parents with ID. Further research comparing these groups will allow clinicians to understand what additional risks are posed to those with ID who are parents over and above the risks attributed to ID in itself.

Although the review did not restrict studies based on language or country, all studies identified were from high-income countries (USA, UK, Sweden, and Canada). However, considering that 80% of the global population with disabilities resides in low- and middle-income countries, it is important that there is research focusing on the mental health needs of women with ID in these countries where healthcare systems and services may be organised differently (World Health Organization, 2011).

## 5. CONCLUSION

Findings from this review suggest that mothers with ID experience disproportionately high rates of mental disorders, particularly when compared with mothers without ID. Routine reproductive healthcare encounters present critical opportunities for prevention and intervention, during which all women should be engaged in discussions about mental health, offered appropriate screening, and provided with individualised, person-centred care. These opportunities are especially salient for women with ID, who continue to encounter barriers within healthcare systems, including stigma and discrimination, which exacerbate existing health inequities. Further research is warranted to elucidate mental health outcomes during reproductive health periods among women with ID and to inform the development of targeted, evidence-based interventions.

## Data Availability

Data availability is not applicable to this article as no new data were created in this study.

## References

Aman, H., Naeem, F., Farooq, S., & Ayub, M. (2016). Prevalence of nonaffective psychosis in intellectually disabled clients: systematic review and meta-analysis. Psychiatric Genetics, 26(4), 145–155. 10.1097/YPG.0000000000000137

American Psychiatric Association. (1994). Diagnostic and statistical manual of mental disorders (IV ed.). Washington, DC: American Psychiatric Association.

American Psychiatric Association. (2013). Diagnostic and statistical manual of mental disorders (5th ed.). Washington, DC: American Psychiatric Association.

Anderson, C., & Cacola, P. (2017). Implications of Preterm Birth for Maternal Mental Health and Infant Development. MCN: The American journal of maternal child nursing, 42(2), 108–114. 10.1097/NMC.0000000000000311

Booth, T., Booth, W., & McConnell, D. (2005). The Prevalence and Outcomes of Care Proceedings Involving Parents with Learning Difficulties in the Family Courts. Journal of Applied Research in Intellectual Disabilities, 18(1), 7–17. 10.1111/j.1468-3148.2004.00204.x

Bowen, E., & Swift, C. (2019). The prevalence and correlates of partner violence used and experienced by adults with intellectual disabilities: A systematic review and call to action. Trauma, Violence, & Abuse, 20(5), 693–705. 10.1177/1524838017728707

Brown, H. K., Cobigo, V., Lunsky, Y., Dennis, C.-L., & Vigod, S. N. (2016). Perinatal Health of Women with Intellectual and Developmental Disabilities and Comorbid Mental Illness. The Canadian Journal of Psychiatry, 61 (11), 714–723. 10.1177/0706743716649188

Brown, H. K., Cobigo, V., Lunsky, Y., & Vigod, S. (2017). Postpartum Acute Care Utilization Among Women with Intellectual and Developmental Disabilities. Journal of Women’s Health, 26(4), 329–337. 10.1089/jwh.2016.5979

Brown, H. K., Vigod, S. N., Fung, K., Chen, S., Guttmann, A., Havercamp, S. M., Parish, S. L., Ray, J. G., & Lunsky, Y. (2022). Perinatal mental illness among women with disabilities: a population-based cohort study. Social psychiatry and psychiatric epidemiology, 57 (11), 2217–2228. 10.1007/s00127-022-02347-2

Carey, I. M., Hosking, F., Harris, T., De Wilde, S., Beighton, C., & Cook, D. G. (2017). An evaluation of the effectiveness of annual health checks and quality of health care for adults with intellectual disability: an observational study using a primary care database. Health Services and Delivery Research, 5(25), 1–158. 10.3310/hsdr05250

Clemente, A. S., Diniz, B. S., Nicolato, R., Kapczinski, F. P., Soares, J. C., Firmo, J. O., & Castro-Costa, É. (2015). Bipolar disorder prevalence: a systematic review and meta-analysis of the literature. Revista Brasileira de Psiquiatria, 37(2), 155–161. 10.1590/1516-4446-2012-1693

Cooper, S.-A., Smiley, E., Morrison, J., Williamson, A., & Allan, L. (2007). Mental ill-health in adults with intellectual disabilities:prevalence and associated factors. British Journal of Psychiatry, 190(1), 27–35. 10.1192/bjp.bp.106.022483

Daveney, J., Hassiotis, A., Katona, C., Matcham, F., & Sen, P. (2019). Ascertainment and Prevalence of Post-Traumatic Stress Disorder (PTSD) in People with Intellectual Disabilities. Journal of Mental Health Research in Intellectual Disabilities, 12(3), 211–233. 10.1080/19315864.2019.1637979

Ehlers-Flint, M. L. (2002). Parenting Perceptions and Social Supports of Mothers with Cognitive Disabilities. Sexuality and Disability, 20(1), 29–51. 10.1023/A:1015282320460

Fairthorne, J., Bourke, J., O’Donnell, M., Wong, K., de Klerk, N., Llewellyn, G., & Leonard, H. (2020). Pregnancy and birth outcomes of mothers with intellectual disability and their infants: Advocacy needed to improve well-being. Disability and Health Journal, 13(2), Article 100871. 10.1016/j.dhjo.2019.100871

Falconer, J. (2018). Removing duplicates from an EndNote library. Library, Archives & Open Research Services Blog. https://blogs.lshtm.ac.uk/library/2018/12/07/removing-duplicates-from-an-endnote-library/

Gaskin, K., & James, H. (2006). Using the Edinburgh Postnatal Depression Scale with learning disabled mothers. Community practitioner : the journal of the Community Practitioners’ & Health Visitors’ Association, 79(12), 392–396.

Heifetz, M., Brown, H. K., Chacra, M. A., Tint, A., Vigod, S., Bluestein, D., & Lunsky, Y. (2019). Mental health challenges and resilience among mothers with intellectual and developmental disabilities. Disability and health journal, 12(4), 602–607. 10.1016/j.dhjo.2019.06.006

Human Rights Act 1998, c. 42. (UK). https://www.legislation.gov.uk/ukpga/1998/42 Equality Act 2010, c. 15. (UK). https://www.legislation.gov.uk/ukpga/2010/15

Hong, Q. N., Pluye, P., Fàbregues, S., Bartlett, G., Boardman, F., Cargo, M., Dagenais, P., Gagnon, M.-P., Griffiths, F., Nicolau, B., O’Cathain, A., Rousseau, M.-C., & Vedel, I. (2018). Mixed methods appraisal tool (MMAT), version 2018. Canadian Intellectual Property Office, Registration of copyright # 1148552.

Javaid, S. F., Hashim, I. J., Hashim, M. J., Stip, E., Samad, M. A., & Ahbabi, A. A. (2023). Epidemiology of anxiety disorders: global burden and sociodemographic associations. Middle East Current Psychiatry, 30, Article 44. 10.1186/s43045-023-00315-3

Lindblad, I., Landgren, V., Gillberg, C., & Fernell, E. (2024). Children born to parents with mild intellectual disability: Register-based follow-up of psychiatric and neurodevelopmental diagnoses and out-of-home placements. Acta Paediatrica, 113(7), 1637–1643. 10.1111/apa.17218

Lo, H. W. J., Poston, L., Wilson, C. A., Sheehan, R., & Sethna, V. (2025). Pregnancy and postnatal outcomes for women with intellectual disability and their infants: A systematic review. Midwifery, 142, Article 104298. 10.1016/j.midw.2025.104298

Lopez-Gonzalez, D. M., & Kopparapu, A. K. (2022). Postpartum care of the new mother. In StatPearls. StatPearls Publishing. https://www.ncbi.nlm.nih.gov/books/NBK565875/

Maïano, C., Coutu, S., Tracey, D., Bouchard, S., Lepage, G., Morin, A. J. S., & Moullec, G. (2018). Prevalence of anxiety and depressive disorders among youth with intellectual disabilities: A systematic review and meta-analysis. Journal of Affective Disorders, 236, 230–242. 10.1016/j.jad.2018.04.029

Maulik, P. K., Mascarenhas, M. N., Mathers, C. D., Dua, T., & Saxena, S. (2011). Prevalence of intellectual disability: a meta-analysis of population-based studies. Research in Developmental Disabilities, 32(2), 419–436. 10.1016/j.ridd.2010.12.018

Mazza, M. G., Rossetti, A., Crespi, G., & Clerici, M. (2020). Prevalence of co-occurring psychiatric disorders in adults and adolescents with intellectual disability: A systematic review and meta-analysis. Journal of Applied Research in Intellectual Disabilities, 33(2), 126–138. 10.1111/jar.12654

McGaw, S., Scully, T., & Pritchard, C. (2010). Predicting the unpredictable? Identifying high-risk versus low-risk parents with intellectual disabilities. Child Abuse & Neglect, 34(9), 699–710. 10.1016/j.chiabu.2010.02.006

McGaw, S., Shaw, T., & Beckley, K. (2007). Prevalence of Psychopathology Across a Service Population of Parents With Intellectual Disabilities and Their Children. Journal of Policy and Practice in Intellectual Disabilities, 4(1), 11–22. 10.1111/j.1741-1130.2006.00093.x

McGowan, J., Sampson, M., Salzwedel, D. M., Cogo, E., Foerster, V., & Lefebvre, C. (2016). PRESS Peer Review of Electronic Search Strategies: 2015 Guideline Statement. Journal of clinical epidemiology, 75, 40–46. 10.1016/j.jclinepi.2016.01.021

Mitra, M., Parish, S. L., Clements, K. M., Cui, X., & Diop, H. (2015). Pregnancy Outcomes Among Women with Intellectual and Developmental Disabilities. American Journal of Preventive Medicine, 48(3), 300–308. 10.1016/j.amepre.2014.09.032

Mueller, B. A., Crane, D., Doody, D. R., Stuart, S. N., & Schiff, M. A. (2019). Pregnancy course, infant outcomes, rehospitalization, and mortality among women with intellectual disability. Disability and health journal, 12(3), 452–459. 10.1016/j.dhjo.2019.01.004

Munshi, S. C., Hoex, E. C. I., Weggelaar-Jansen, A. M., Knijff, E. M., van der Wilk, E. C., Steegers, E. A. P., & Bijma, H. H. (2025). Integrated care for multi-domain vulnerability during pregnancy: a retrospective cohort study. Archives of women’s mental health, 28(4), 699–709. 10.1007/s00737-024-01554-x

Oeseburg, B., Dijkstra, G. J., Groothoff, J. W., Reijneveld, S. A., & Jansen, D. E. M. C. (2011). Prevalence of Chronic Health Conditions in Children With Intellectual Disability: A Systematic Literature Review. Intellectual and Developmental Disabilities, 49(2), 59–85. 10.1352/1934-9556-49.2.59

Parish, S. L., Mitra, M., Son, E., Bonardi, A., Swoboda, P. T., & Igdalsky, L. (2015). Pregnancy Outcomes Among U.S. Women With Intellectual and Developmental Disabilities. American Journal on Intellectual and Developmental Disabilities, 120(5), 433–443. 10.1352/1944-7558-120.5.433

Powell, R. M., Parish, S. L., Mitra, M., Waterstone, M., & Fournier, S. (2024). Child welfare system inequities experienced by disabled parents: towards a conceptual framework. Disability & Society, 39(2), 291–318. 10.1080/09687599.2022.2071675

Rahman, T., & Lauriello, J. (2016). Schizophrenia: An Overview. Focus (American Psychiatric Publishing), 14(3), 300–307. 10.1176/appi.focus.20160006

Saldanha, I. J., Adam, G. P., Kanaan, G., Zahradnik, M. L., Steele, D. W., Danilack, V. A., Peahl, A. F., Chen, K. K., Stuebe, A. M., & Balk, E. M. (2023). Postpartum Care up to 1 Year After Pregnancy: A Systematic Review and Meta-Analysis. Agency for Healthcare Research and Quality (US). 10.23970/AHRQEPCCER261

Shea, L., Sadowsky, M., Tao, S., Rast, J., Schendel, D., Chesnokova, A., & Headen, I. (2024). Perinatal and Postpartum Health Among People With Intellectual and Developmental Disabilities. JAMA network open, 7(9), e2428067. 10.1001/jamanetworkopen.2024.28067

Sterling, J. D. (1998). Determinants of parenting in women with mental retardation (Order No. 9908035). Available from ProQuest Dissertations & Theses Global. (304468501). https://www.proquest.com/dissertations-theses/determinants-parenting-women-with-mental/docview/304468501/se-2

Symonds, J., Abbott, D., & Dugdale, D. (2021). “Someone will come in and say I’m doing it wrong.” The perspectives of fathers with learning disabilities in England. British Journal of Learning Disabilities, 49(1), 23–33. 10.1111/bld.12351

Tarasoff, L. A., Lunsky, Y., Chen, S., Guttmann, A., Havercamp, S. M., Parish, S. L., Vigod, S. N., Carty, A., & Brown, H. K. (2020). Preconception Health Characteristics of Women with Disabilities in Ontario: A Population-Based, Cross-Sectional Study. Journal of women’s health, 29(12), 1564–1575. 10.1089/jwh.2019.8273

Turney, D., Tarleton, B., & Tilbury, N. (2018). Supporting parents who have learning disabilities: Strategic Briefing: Dartington: Research in Practice. https://www.researchinpractice.org.uk/all/publications/2018/april/supporting-parents-who-have-learning-disabilities-strategic-briefing-2018/

Tymchuk, A. (1993). Symptoms of psychopathology in mothers with mental handicap. Mental Handicap Research, 6(1), 18–35. 10.1111/j.1468-3148.1993.tb00048.x

Tymchuk, A. J. (1994). Depression symptomatology in mothers with mild intellectual disability: An exploratory study. Australia and New Zealand Journal of Developmental Disabilities, 19(2), 111–119. 10.1080/07263869400035151

Varcin, K. J., Herniman, S. E., Lin, A., Chen, Y., Perry, Y., Pugh, C., Chisholm, K., Whitehouse, A. J. O., & Wood, S. J. (2022). Occurrence of psychosis and bipolar disorder in adults with autism: A systematic review and meta-analysis. Neuroscience & Biobehavioral Reviews, 134, Article 104543. 10.1016/j.neubiorev.2022.104543

Walton-Allen, N. (1993). Psychological distress and parenting by mothers with mental retardation (Order No. NN86365). Available from ProQuest Dissertations & Theses Global. (304083774). https://www.proquest.com/dissertations-theses/psychological-distress-parenting-mothers-with/docview/304083774/se-2

White, A., Sheehan, R., Ding, J., Roberts, C., Magill, N., Keagan-Bull, R., Carter, B., Chauhan, U., Tuffrey-Wijne, I., & Strydom, A. (2023). Learning from lives and deaths – People with a learning disability and autistic people (LeDeR) report for 2022. King’s College London. https://www.kcl.ac.uk/research/leder

World Health Organization. (2004). International statistical classification of diseases and related health problems *(10th rev.,* 2nd *ed.)*. World Health Organization. https://apps.who.int/iris/handle/10665/42980

World Health Organization. (2011). World report on disability. Geneva: World Health Organization. Available from: https://www.ncbi.nlm.nih.gov/books/NBK304079/

World Health Organization. (2022). International classification of diseases for mortality and morbidity statistics (11th rev.). World Health Organization. https://icd.who.int/en

World Health Organization. (2025). Depressive disorder (depression). World Health Organization. https://www.who.int/news-room/fact-sheets/detail/depression

