## Supplementary Materials for "Prevalence of Mental Disorders in Parents with Intellectual Disabilities: Systematic Review and Meta-Analysis"

**S1: Database Search Strategy** The final search strategy for MEDLINE, APA PsycINFO and EMBASE are shown. Search terms were adjusted between databases due to different MeSH/Emtree/Index terms and syntax between databases.

**MEDLINE (Ovid) Search Strategy**

Within each concept below, every search term will be connected using the Boolean operator “OR” and all concepts are linked using the Boolean operator “AND”.

**Concept 1:**

Intellectual Disability/

Fragile X Syndrome/

Down Syndrome/

(intellectual* adj4 (disab* or disorder* or difficult*)).ti,ab,kf

("learning disabilit*" or "mental* retard*" or "learning difficult*" or "mental* handicap*" or “developmental disabilit* or "fragile x" or "martin-bell" or fxs or "Fraxa" or "marker x syndrome" or "down* syndrome" or "trisomy 21*”).ti,ab,kf.

**Concept 2:**

Parents/

Pregnancy/

Postpartum period/

Perinatal Care/

Postnatal Care/

(parent* or mother* or father* or pregnan* or perinatal or postpartum or postnatal or prenatal or antenatal or peri-natal or post-partum or post-natal or pre-natal or ante-natal).ti,ab,kf

**Concept 3:**

Mental Disorders/

(psychiatri* or hypochondriasis or “conversion disorder*” or “obsessive-compulsive” or “obsessive compulsive” or OCD or “adjustment disorder*” or PTSD or “post traumatic stress disorder*” or “post-traumatic stress disorder*” or “eat* disorder*” or “feeding disorder*” or “binge eat*” or anorex* or bulimi* or delusion* or bipolar or BPAD or mania or cyclothymi* or psychos?s or psychotic or schizo* or catatoni* or “somat* disorder” or depression or depressive or dysthymi* or anxiety or phobi* or panic or “personality disorder*” or “paraphilic disorder*” or “impulse control disorder*”).ti,kf,ab.

(mental adj2 (condition* or disorder* or ill* or diagnos?s or problem* or health*)).ti, kf, ab

(affective adj2 (disorder* or problem*)).ti, kf, ab

(mood adj2 (disorder* or problem*)).ti, kf, ab

(compulsive adj2 (eat* or vomit* or purg*).ti, kf, ab

(substance adj2 (dependenc* or addiction or abuse or misuse or disorder*)).ti, kf, ab

(dissociative adj2 disorder*).ti,ab,kf.

**EMBASE Search Strategy**

Emtree terms based on those in MEDLINE strategy, with the addition of Concept 2 containing additional Emtree term not available in MEDLINE: pregnant woman/

Free text-text search as per MEDLINE strategy.

**PsycInfo Search Strategy**

Index terms based on those in MEDLINE (Ovid) strategy.

Free text searches as per MEDLINE (Ovid) strategy, limited to title, abstract and keywords using .ti, ab id.

**S2: Articles excluded** Details of one report not retrieved and 37 articles excluded during full-text screening. As shown, common reasons for exclusion were: studies focused on “Intellectual and Developmental Disabilities” (IDD) did not give separate results for those with “Intellectual Disability” (ID); diagnoses of intellectual disability were not made using formal diagnostic criteria; and mental disorder outcomes were not reported.

|  | **Reports not retrieved** | **Details** |  |
| --- | --- | --- | --- |
|  | Mentally Disabled Women Who Have a Child Anxiety Levels and Psychosocial Support Needs Determination; S Mert Boğa, N Köşgeroğlu, FD Sayıner, N Özerdoğan, A Açıkgöz    Abstracts from the 4th World Congress on Women’s Mental Health. *Arch Womens Ment Health* 14 (Suppl 2), 89–163 (2011). https://doi.org/10.1007/s00737-011-0238-y | Conference abstract – unable to identify corresponding published article |  |
|  | **Article excluded at full-text screening** | **Primary reason for Exclusion** |  |
| 1 | Brown, H. K., Cobigo, V., Lunsky, Y., Dennis, C.-L., & Vigod, S. (2016). Perinatal Health of Women with Intellectual and Developmental Disabilities and Comorbid Mental Illness. Canadian journal of psychiatry. Revue canadienne de psychiatrie, 61(11), 714-723. https://doi.org/https://dx.doi.org/10.1177/0706743716649188 | No separate outcome data for those with ID only | From database search |
| 2 | Brown, H. K., Lunsky, Y., Wilton, A. S., Cobigo, V., & Vigod, S. N. (2016). Pregnancy in Women With Intellectual and Developmental Disabilities. Journal of obstetrics and gynaecology Canada : JOGC = Journal d'obstetrique et gynecologie du Canada : JOGC, 38(1), 9-16. https://doi.org/https://dx.doi.org/10.1016/j.jogc.2015.10.004 | No separate outcome data for those with ID only | From database search |
| 3 | Brown, H. K., Potvin, L. A., Lunsky, Y., & Vigod, S. N. (2018). Maternal Intellectual or Developmental Disability and Newborn Discharge to Protective Services. Pediatrics, 142(6). https://doi.org/https://dx.doi.org/10.1542/peds.2018-1416 | No separate outcome data for those with ID only | From database search |
| 4 | Brown, H. K., Ray, J. G., Chen, S., Guttmann, A., Havercamp, S. M., Parish, S., Vigod, S. N., Tarasoff, L. A., & Lunsky, Y. (2021). Association of Preexisting Disability With Severe Maternal Morbidity or Mortality in Ontario, Canada. JAMA network open, 4(2), e2034993. https://doi.org/https://dx.doi.org/10.1001/jamanetworkopen.2020.34993 | No separate outcome data for those with ID only | From database search |
| 5 | Brown, H. K., Varner, C., Ray, J. G., Scime, N. V., Fung, K., Guttmann, A., Havercamp, S. M., Vigod, S. N., & Lunsky, Y. (2023). Comparison of Emergency Department Use Between Pregnant People With and Without Disabilities in Ontario, Canada. JAMA network open, 6(8), e2327185. https://doi.org/https://dx.doi.org/10.1001/jamanetworkopen.2023.27185 | No separate outcome data for those with ID only | From database search |
| 6 | Brown, H. K., Vigod, S. N., Fung, K., Chen, S., Guttmann, A., Havercamp, S. M., Parish, S. L., Ray, J. G., & Lunsky, Y. (2022). Perinatal mental illness among women with disabilities: a population-based cohort study. Social psychiatry and psychiatric epidemiology, 57(11), 2217-2228. https://doi.org/https://dx.doi.org/10.1007/s00127-022-02347-2 | No separate outcome data for those with ID only | From database search |
| 7 | Cook, V. H. (2009). Does depression and low literacy in mothers affect their interactions with their 4-month old infants? Dissertation Abstracts International Section A: Humanities and Social Sciences, 70(3), 840. | Population - Available measures of very low literacy were used as “proxy” measures for intellectual disability. However, lowest IQ group mean above 70 indicates unlikely ID sample. | From database search |
| 8 | Edwards, N. M., Lieberman-Betz, R., & Wiegand, S. (2023). Parents with intellectual disability and mental health conditions: Early intervention providers' perceptions. Journal of intellectual & developmental disability, 48(3), 225-237. https://doi.org/https://dx.doi.org/10.3109/13668250.2022.2112530 | Population not ID | From database search |
| 9 | Emerson, E., & Brigham, P. (2013). Health behaviours and mental health status of parents with intellectual disabilities: cross sectional study. Public health, 127(12), 1111-1116. https://doi.org/https://dx.doi.org/10.1016/j.puhe.2013.10.001 | Population – ID self-reported by the item ‘parent(s) have learning difficulties’. Data was collected at level of households (rather than individuals). | From database search |
| 10 | Hindmarsh, G., Llewellyn, G., & Emerson, E. (2015). Mothers with intellectual impairment and their 9-month-old infants. Journal of Intellectual Disability Research, 59(6), 541-550. https://ovidsp.ovid.com/ovidweb.cgi?T=JS&CSC=Y&NEWS=N&PAGE=fulltext&D=med12&DO=10.1111%2fjir.12159 | ID diagnosis not based on clinical diagnostic criteria – No mothers reported having a disability coded in the ICD blocks F70–79. Majority identified as having “intellectual impairment” based on poor literacy, numeracy and low educational attainment. | From database search |
| 11 | Horner-Johnson, W., Garg, B., Darney, B. G., Biel, F. M., & Caughey, A. B. (2022). Severe maternal morbidity and other perinatal complications among women with physical, sensory, or intellectual and developmental disabilities. Paediatric and perinatal epidemiology, 36(5), 759-768. https://doi.org/https://dx.doi.org/10.1111/ppe.12873 | No separate outcome data for those with ID only | From database search |
| 12 | Jha, S., Khanna, A., & Khanna, S. K. (2015). Mother Child Unit (MCU) of a tertiary care institution: A first cry from India. Asian Journal of Psychiatry, 16, 72. | Population - Only 33% of sample had ID and their MH results are not given separately | From database search |
| 13 | Jones, K. B., Taylor, I. K., Schwab, T., King, C., Okoye, G., & Kim, J. (2025). Factors Associated with Adverse Birth Outcomes in Women with an Intellectual or Other Developmental Disability. Healthcare (Basel, Switzerland), 13(7). https://doi.org/https://dx.doi.org/10.3390/healthcare13070780 | No separate outcome data for those with ID only | From database search |
| 14 | Kassee, C., Lunsky, Y., Patrikar, A., & Brown, H. K. (2023). Impact of social-, health-, and disability-related factors on pregnancy outcomes in women with intellectual and developmental disabilities: A population-based latent class analysis. Disability and health journal, 16(2), 101426. https://dx.doi.org/10.1016/j.dhjo.2022.101426 | No separate outcome data for those with ID only | From database search |
| 15 | Laxova, R., Gilderdale, S., & Ridler, M. A. (1973). An aetiological study of fifty-three female patients from a subnormality hospital and of their offspring. Journal of Mental Deficiency Research, 17(3), 193-225. | Patients with psychiatric disorders do not meet ID criteria for IQ | From database search |
| 16 | Lewis, D. O., Klerman, L. V., Jekel, J. F., & Currie, J. B. (1973). Experiences with psychiatric services in a program for pregnant school-age girls. Social Psychiatry, 8(1), 16-25. | Population - the 3 who refused to be seen potentially ID as mean IQ 63 - other groups all above 70. However, there is no separate breakdown of results for the group with potential ID. | From database search |
| 17 | Lima, F., O'Donnell, M., Bourke, J., Wolff, B., Gibberd, A., Llewellyn, G., & Leonard, H. (2022). Child protection involvement of children of mothers with intellectual disability. Child abuse & neglect, 126, 105515. https://doi.org/https://dx.doi.org/10.1016/j.chiabu.2022.105515 | Outcome is maternal mental health contact and maternal substance use contact rather than diagnosis. | From database search |
| 18 | Matone, M., Minkovitz, C., Quarshie, W., & Rubin, D. M. (2016). Chronic disease prevalence and discontinuation of medications among young mothers with a relationship to the child welfare system. Children and Youth Services Review, 64, 66-72. | Population - Percent with ID in samples is between 0.3 - 9 percent. | From database search |
| 19 | McConnell, D., Dalziel, A., Llewellyn, G., Laidlaw, K., & Hindmarsh, G. (2009). Strengthening the social relationships of mothers with learning difficulties. British Journal of Learning Disabilities, 37(1), 66-75. | Population - Unclear proportion with formal diagnosis of ID rather than “learning difficulties” | From database search |
| 20 | McConnell, D., Mayes, R., & Llewellyn, G. (2008). Pre-partum distress in women with intellectual disabilities. Journal of intellectual & developmental disability, 33(2), 177-183. https://doi.org/https://dx.doi.org/10.1080/13668250802007903 | Population – Diagnosis of ID not based on formal clinical diagnostic criteria | From database search |
| 21 | Munshi, S. C., Hoex, E. C. I., Weggelaar-Jansen, A. M., Knijff, E. M., van der Wilk, E. C., Steegers, E. A. P., & Bijma, H. H. (2025). Integrated care for multi-domain vulnerability during pregnancy: a retrospective cohort study. Archives of women's mental health. https://dx.doi.org/10.1007/s00737-024-01554-x | Population - 35/91 (38%) of women with ID were identified through a formal assessment of IQ and 56/91 (56%) by clinical suspicion not formal diagnosis. | From database search |
| 22 | Murthy, G. V. S., John, N., Sagar, J., Shamanna, B. R., Noe, C., Soji, F., Mani, S., Pant, H. B., Allagh, K., & Kamalakannan, S. (2014). Reproductive health of women with and without disabilities in South India, the SIDE study (South India Disability Evidence) study: A case control study. BMC Women's Health, 14. | No separate analysis for ID. | From database search |
| 23 | Sapkota, D., Ogilvie, J., Dennison, S., Thompson, C., & Allard, T. (2024). Prevalence of mental disorders among Australian females: Comparison according to motherhood status using Australian birth cohort data. Archives of women's mental health, 27, 625-635. https://ovidsp.ovid.com/ovidweb.cgi?T=JS&CSC=Y&NEWS=N&PAGE=fulltext&D=emed26&DO=10.1007%2fs00737-024-01444-2 | No separate data for mothers with ID and another psychiatric comorbidity (arbitrated) | From database search |
| 24 | Tebes, J. K., Kaufman, J. S., Adnopoz, J., & Racusin, G. (2001). Resilience and family psychosocial processes among children of parents with serious mental disorders. Journal of Child and Family Studies, 10(1), 115-136. | Population – no evidence of ID | From database search |
| 25 | Warner, N., Scourfield, J., Cannings-John, R., Rouquette, O. Y., Lee, A., Vaughan, R., Broadhurst, K., & John, A. (2024). Parental risk factors and children entering out-of-home care: The effects of cumulative risk and parent's sex. Children and Youth Services Review, 160, 1-13. | Population - Frequency of ID in cohort is 0.9% - there is not an individual break down of their Mental Health diagnoses | From database search |
| 26 | Watkeys, O. J., O'Hare, K., Dean, K., Laurens, K. R., Harris, F., Carr, V. J., & Green, M. J. (2023). Early childhood developmental vulnerability associated with parental mental disorder comorbidity. Australian and New Zealand Journal of Psychiatry, 57, 1117-1129. | No separate data for ID | From database search |
| 27 | Zhou, M., Larsson, H., D'Onofrio, B. M., Landen, M., Lichtenstein, P., & Pettersson, E. (2023). Intergenerational Transmission of Psychiatric Conditions and Psychiatric, Behavioral, and Psychosocial Outcomes in Offspring. JAMA network open, 6(12), e2348439. https://doi.org/https://dx.doi.org/10.1001/jamanetworkopen.2023.48439 | Population – no ID in parents | From database search |
| 28 | Tarasoff, L. A., Lunsky, Y., Chen, S., Guttmann, A., Havercamp, S. M., Parish, S. L., Vigod, S. N., Carty, A., & Brown, H. K. (2020). Preconception Health Characteristics of Women with Disabilities in Ontario: A Population-Based, Cross-Sectional Study. *Journal of women's health (2002)*, *29*(12), 1564–1575. https://doi.org/10.1089/jwh.2019.8273 | Population – preconception rather than during pregnancy/postpartum/parenthood | From Conference Abstract search |
| 29 | Rosenberg, S., & McTate, G. (1990). Intellectually handicapped mothers: Problems an d prospects. Children Today. 1 1 .1-9. | Outcome – no mental health outcomes | From reference search |
| 30 | Cadamy, P. (2002). A description of the needs of mothers enrolled in a program for at -risk parents (Order No. 3054057). Available from ProQuest Dissertations & Theses Global. (305550695). Retrieved from <https://www.proquest.com/dissertations-theses/description-needs-mothers-enrolled-program-at/docview/305550695/se-2> | Population - Participants from “Project Employ”, no indication requiring ID. IQ levels from WASI 70 – 101 indicate unlikely ID | From grey literature search |
| 31 | Brown, H. K., Kirkham, Y. A., Cobigo, V., Lunsky, Y., & Vigod, S. N. (2016). Labour and delivery interventions in women with intellectual and developmental disabilities: a population-based cohort study. *J Epidemiol Community Health*, *70*(3), 238-244. | IDD no separate ID data | From citation search |
| 32 | Grant, C., Lunsky, Y., Guttmann, A., Vigod, S. N., Sharpe, I., Fung, K., & Brown, H. K. (2024). Maternal disability and newborn discharge to social services: a population-based study. *International Journal of Population Data Science*, *9*(2), 2396. | IDD no separate ID data | From citation search |
| 33 | Feldman, M. A., Varghese, J., Ramsay, J., & Rajska, D. (2002). Relationships between Social Support, Stress and Mother--Child Interactions in Mothers with Intellectual Disabilities. *Journal of Applied Research in Intellectual Disabilities*, *15*(4). | No mental disorder outcome | From grey literature |
| 34 | Elvish, J., Hames, A., English, S., & Wills, C. (2006). Parents with learning disabilities: an audit of referrals made to a learning disability team. *Tizard Learning Disability Review*, *11*(2), 26-33. | No mental disorder outcome | From grey literature |
| 35 | Malouf, R., Henderson, J., & Redshaw, M. (2017). Access and quality of maternity care for disabled women during pregnancy, birth and the postnatal period in England: data from a national survey. *BMJ open*, *7*(7), e016757. | No mental disorder outcome | From grey literature |
| 36 | Murphy, G. and Feldman, M.A. (2002), Parents with Intellectual Disabilities. Journal of Applied Research in Intellectual Disabilities, 15: 281-284. <https://doi.org/10.1046/j.1468-3148.2002.00139.x> | No mental disorder outcome | From grey literature |
| 37 | Stenfert Kroese, B., Hussein, H., Clifford, C., & Ahmed, N. (2002). Social support networks and psychological well‐being of mothers with intellectual disabilities. *Journal of Applied Research in Intellectual Disabilities*, *15*(4), 324-340. | No mental disorder outcome | From grey literature |

**S3: Detailed MMAT Results and Justifications** The Mixed Methods Appraisal Tool (MMAT; Hong et al. (2018)), evaluates studies using two universal screening questions and five design-specific criteria, rated as “Yes,” “No,” or “Can’t tell,” to ensure transparent appraisal across study types. In order to aid comparisons across studies, a final judgement was given by each assessor depending on whether the study met the criteria on all 5 questions (low risk of bias), 4 questions (moderate risk of bias), or ≤3 questions (high risk of bias). Any missing or unclear data was judged to reduce the overall risk of bias of the paper.

| **MMAT Category** | **Study**  **First author, year** | **S1** | **S2** | **Q1** | **Q2** | **Q3** | **Q4** | **Q5** | **Risk of Bias Judgement** |
| --- | --- | --- | --- | --- | --- | --- | --- | --- | --- |
|  |  | **Are there clear research questions?** | **Do the collected data allow to address the research questions?** | **Is the sampling strategy relevant to address the research question?** | **Is the sample representative of the target population?** | **Are the measurements appropriate?** | **Is the risk of nonresponse bias low?** | **Is the statistical analysis appropriate to answer the research question?** |  |
| **Quantitative Descriptive** | **Sterling 1999** | Yes | Yes | Yes | No | Yes | Can’t tell | Yes | High |
|  | **Justification** | The sample included mothers with intellectual disabilities. There is complete data for CES-D from all mothers identified for study. Analyses done as part of wider study was appropriate and prevalence for depression obtained from descriptive information.  The study used CES-D which was developed with middle class caucasians with reasonable education. Although the author adapted and piloted it before data collection, the CES-D measures the symptoms of depression on a continuum and is not meant to diagnose clinical depression  The women in the study were identified by Regional Centre case managers. It is not clear how many of the total women they referred refused to participate or could not be contacted.  A convenience sample was drawn from 2 regional centres, thereby excluding women not engaged with these services. Also mothers with a diagnosis of the more severe forms of psychopathology such as psychotic processes and /or personality disorders, which are known to interfere with parenting adequacy, were not included. This reduced the representativeness and generalisability of findings to the populations who are most engaged with ID services. | | | | | | | |
|  | **McGaw 2007** | Yes | Yes | Yes | No | Yes | No | Yes | High |
|  | **Justification** | Efforts were made to ensure that a cross-section of families were included in the sample, in terms of their parental functioning and risk status. The study used WAIS to measure parental IQ and mini PAS-ADD to measure psychopathology - Studies of this tool’s reliability have been demonstrated in its complete form, although in this study the short-form checklist was used. The study used appropriate analyses to examine associations.  The sample represented 21% of families referred to the parenting service. The remaining 79% of nonparticipating families from the service population either did not meet the eligibility criteria (66%), declined to take part in the study (8%), or the family was engaged in child care proceedings and were at risk of their child being removed into care out-of-home (5%). Eligibility included that the parents having one or more children aged 5 years–18 years who were residing in the family home. This limits generalizability of the sample. Additionally, during the period of the research program, one family disengaged from the project and five of the 65 interview sessions were missed by participant parents. | | | | | | | |
|  | **Walton-Allen 1993** | Yes | Yes | Yes | No | Yes | No | Yes | High |
|  | **Justification** | Social service agencies, who are often involved with mothers with ID, identified mothers for the study. The study explores the prevalence of depressive symptoms and parenting stress in mothers with intellectual disabilities using BDI, which has been widely used even with individuals with mild intellectual disabilities. The Wechsler Adult Intelligence Scale - Revised (WAIS-R; Wechsler, 1981) is a valid method which was used to obtain a measure of intellectual functioning. Social service agencies, often involved with mothers with ID, referred mothers. Of 40 included in study, there were responses from all 40. Descriptive statistics were appropriate. Cut-off criteria from tool manuals were correctly applied.  Social service agencies identified 57 eligible mothers (of these 5 refused, 12 not eligible). Due to the small numbers in the study, the refusals may bias the result.  One of eligibility criteria was that mother has custody of her child. Only one mother was Asian and the rest Caucasian, and the sample only includes mothers known to social services. For these reasons the sample may not be representative and generalisability is limited. | | | | | | | |
|  | **McGaw 2010** | Yes | Yes | Yes | No | Yes | Yes | Yes | Moderate |
|  | **Justification** | This was a secondary data analysis of service users who had consented to data collection. It is not explicit whether any individuals declined to consent but “participants comprised a population from a UK based NHS Special Parenting Service” implies the whole cohort was included.  Formal assessments and diagnosis were made based on DSM-IV, AAMR, and ICD 10 classifications and psychometric testing were used as confirmatory evidence to substantiate parent and caseworker reports. The analyses were appropriate for the goals of the study. There is no indication of missing data from those who were included.  Participants comprised a population of adults who had received advice, guidance and support over the period 1999 to 2004 from a UK-based NHS Special Parenting Service based in a largely rural area across the South West of the UK, making generalizability limited. | | | | | | | |
|  | **Gaskin 2006** | Yes | Yes | Yes | No | Yes | Can’t tell | Yes | High |
|  | **Justification** | Health visitors, midwives, community nurses, community consultant paediatricians, social workers and family support workers were approached and asked to identify mothers with ID.  The EDPS and Wechsler Abbreviated Scale of Intelligence (WASI) are validated tools which were used. For all mothers who took part there appears to be a complete EDPS. Analyses were appropriate for the study.  There is no information about who declined to participate or who was not reached  The sample was small (13 women, 15 data sets) and mostly White British, limiting generalizability. There is no indication of the number who could not be contacted or declined to participate. | | | | | | | |
|  | **Heifetz 2019** | Yes | Yes | Yes | No | Yes | Can’t tell | Yes | High |
|  | **Justification** | In this “qualitative study” the authors collect quantitative data to describe the sample, which were not linked to the qualitative findings. The qualitative findings have not been extracted for this systematic review and thus assessing quality using the “qualitative” criteria was deemed not relevant. The lack of integration between qualitative and quantitive components means the study cannot be assessed using “mixed methods criteria”. Thus the “quantitative descriptive” MMAT category was deemed most appropriate.  The study used convenience sampling from an urban agency providing parenting support to mothers with IDD. Women were eligible to participate if they had previously been diagnosed with IDD (IQ below 70 and below average adaptive skills).  Depression was measured with the Glasgow Depression Scale—validated for adults with IDD, which showed an acceptable internal concistency in this sampe (Cronbach’s α = .756). Quantitative data were appropriately summarized for sample description.  The small sample, recruited from one service agency, currently receiving specialized services; with custody of their children and all English speaking, limits the generalisability of the findings.  The study also did not report the number of women who refused to participate in the study or did not respond. | | | | | | | |
|  |  | **Are there clear research questions?** | **Do the collected data allow to address the research questions?** | **Are the participants representative of the target population?** | **Are measurements appropriate regarding both the outcome and exposure?** | **Are there complete outcome data?** | **Are the confounders accounted for in the design and analysis?** | **During the study period, were the exposure and the outcome identified and classified as intended?** |  |
| **Quantitative non-randomized** | **Tymchuk 1994** | Yes | Yes | No | Yes | Yes | Yes | Yes | Moderate |
|  | **Justification** | The WAIS used to confirm IQ and The Beck Depression Inventory (BDI) used to measure depression in adults. This is a validated, appropriate tool for measuring depression in adults, including those with mild ID. Outcome data (BDI scores) were provided for all participants. The analysis explored some confounders.  The study includes mothers participating in a community and university hospital based programme designed to provide clinical services to mothers with ID. The sample is small and non-random. Thus results cannot be generalized beyond this group. The contrast group had children in daycare groups in the same area and volunteered to participate. | | | | | | | |
|  | **Lindblad 2024** | Yes | Yes | No | Yes | Yes | No | Yes | Moderate |
|  | **Justification** | The original cohort was derived from a total population of 6397 children living in a Swedish suburban municipality and consisted of 82 children with MID as defined in DSM-IV (American Psychiatric Association 1994), diagnosed in childhood, verified by educational and health service records. 78 were part of a follow up study, of whom 31 had become parents (15 mothers and 16 fathers) and the non-parents were the comparison group. This indicates a small number lost to follow-up and, there appears to be no other missing data.  There is no evidence of adjustment for important potential confounders (e.g., parental comorbidities, socioeconomic status).  Diagnoses were from the national patient register (NPR) - derived from doctor's visits in specialist care. Although the strength of a national register is acknowledged by the moderate rating, this register does not cover primary care visits to primary care or allied health professionals. | | | | | | | |
|  | **Shea 2024** | Yes | Yes | No | Yes | Yes | Yes | Yes | Moderate |
|  | **Justification** | All Medicaid enrolees were included in the study. ID was identfied by at least 1 inpatient or 2 other claims associated with an ID diagnosis and MH conditions by ICD9 Diagnosis codes.  All co-occurring mental health conditions were compared across study groups in bivariate multivariable analyses. The latter comprised calculation of odds ratios for each condition comparing the group with IDD with the group without IDD, adjusting for age at delivery, year of delivery, race and ethnicity, and state of residence. Individuals with no race or ethnicity information were included in a separate “missing” category in both descriptive statistics and models. There was no indication of other missing data,  Unfortunately, the division of the groups for analysis meant that the ID group did not include anyone with autism (even comorbid autism), which is often a significant proportion of those with ID. Data was based on Medicaid claims, thus limiting generalizability to people with IDD with private health insurance. However, people with ID are often of lower socioeconomic status and possibly more likely to use Medicaid. | | | | | | | |
|  | **Tymchuk 1993** | Yes | Yes | No | No | Yes | Yes | Yes | High |
|  | **Justification** | The study includes mothers with ID, with WAIS used to confirm IQ. All 31 mothers, along with comparison groups, completed the assessment. The results appear to have complete data. The mothers in the contrast group were reportedly similar to those with ID on all variables except the number of children.  The mothers in the study were participating in a community and university hospital based programme. The sample is small, non-random and results cannot be generalized beyond this group.  Additionally, the self-report version of the Psychopathology Instrument for Mentally Retarded Adults (PIMRA) was used to assess psychopathology. There are concerns that no decision rules have been developed regarding just how many symptoms presented over what periods of time actually mean that a person with ID has a particular emotional disorder. Also, while the authors suggested that the items were simplified in order to be comprehended by people with mental handicap, there is no description of how that simplification was achieved or of the testing of the items to ensure that they are comprehensible by persons of varying levels of intellectual ability. In addition, there is no indication of which items have been removed in order to make it a 56 item scale and the order in which those items appear. The PIMRA has seen only limited use and reportedly suffers from methodological problems stemming from the manner in which it was constructed. | | | | | | | |

**Meta-Analyses**

**S4a: Forest plot showing 6 studies providing prevalence figures for Depression** The majority of studies provided prevalence data for depression. A meta-analysis of all of these studies showed substantial heterogeneity (I^2 =^ 86.73%, p<0.001).

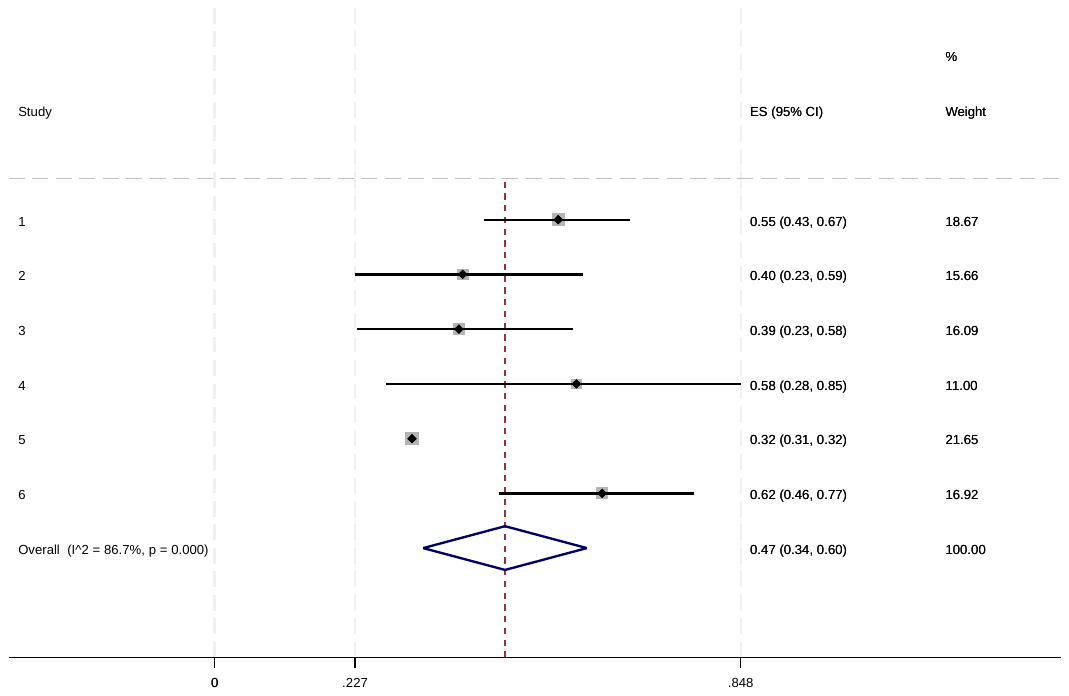

**S4b: Forest plot showing studies providing prevalence figures for Anxiety** The pooled random effect size for studies with anxiety prevalence figures was 0.26 (95% CI 0.26 to 0.26) with no variation attributable to heterogeneity (I^2^ = 0.00%, p=0.888). This indicates a consistent estimate across studies, suggesting that approximately one-quarter of parents experienced anxiety.

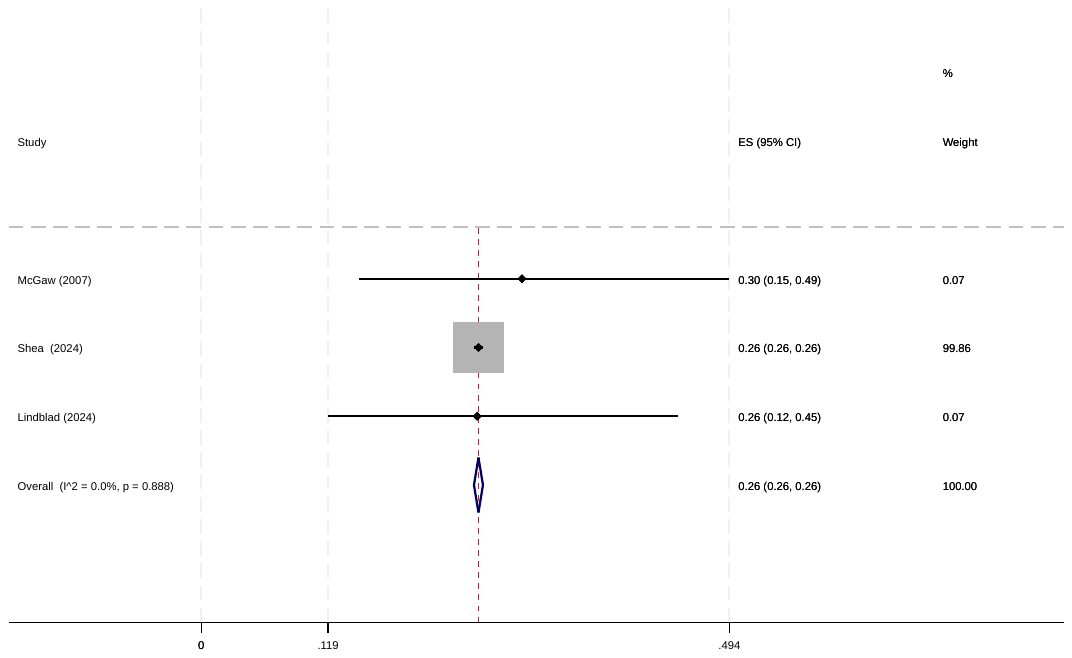

**S4c: Forest plot showing studies providing prevalence figures for Psychosis** The pooled random effect size for studies with psychosis prevalence was 0.09 (95% CI 0.03 to 0.14) with moderate variation attributable to heterogeneity (I^2^ = 61.55%, p0.074). This suggests that 9% of participants experienced psychosis, though the prevalence estimates varied moderately between studies.

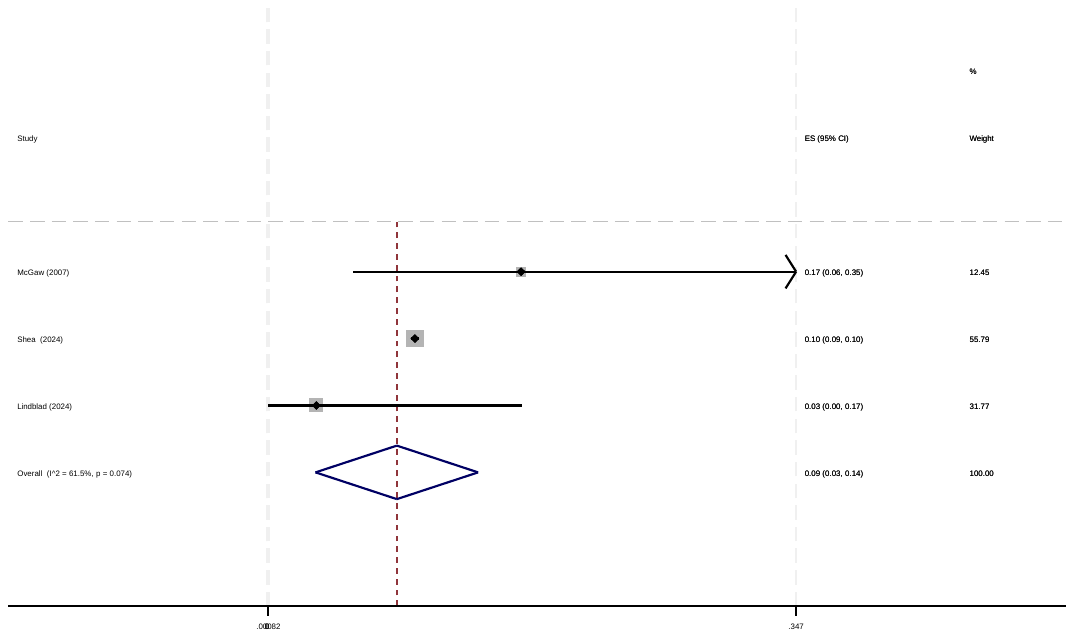
